# Measuring the Impact of Exposure to COVID-19 Vaccine Misinformation on Vaccine Intent in the UK and US

**DOI:** 10.1101/2020.10.22.20217513

**Authors:** S. Loomba, A. de Figueiredo, S. J. Piatek, K. de Graaf, H. J. Larson

## Abstract

The successful development and widespread acceptance of a SARS-CoV-2 vaccine will be a major step in fighting the pandemic, yet obtaining high uptake will be a challenging task, worsened by online misinformation. To help inform successful COVID-19 vaccination campaigns in the UK and US, we conducted a survey to quantify how online misinformation impacts COVID-19 vaccine uptake intent and identify socio-economic groups that are most at-risk of non-vaccination and most susceptible to online misinformation. Here, we report findings from nationally representative surveys in the UK and the US conducted in September 2020. We show that recent misinformation around a COVID-19 vaccine induces a fall in vaccination intent among those who would otherwise “definitely” vaccinate by 6.4 (3.8, 9.0) percentages points in the UK and 2.4 (0.1, 5.0) in the US, with larger decreases found in intent to vaccinate to protect others. We find evidence that socio-econo-demographic, political, and trust factors are associated with low intent to vaccinate *and* susceptibility to misinformation: notably, older age groups in the US are more susceptible to misinformation. We find evidence that scientific-sounding misinformation relating to COVID-19 and vaccines COVID-19 vaccine misinformation lowers vaccination intent, while corresponding factual information does not. These findings reveal how recent COVID-19 misinformation can impact vaccination rates and suggest pathways to robust messaging campaigns.

---

The spread of the novel severe acute respiratory syndrome coronavirus 2 (SARS-CoV-2) that causes coronavirus disease 2019 (COVID-19) has resulted in an unprecedented global public health and economic burden (Andersen 2020; Zhou 2020). Developing a COVID-19 vaccine will be a major step in fighting the disease—which was declared a pandemic by the World Health Organization on 11 March 2020 (WHO 2020). In order for a novel COVID-19 vaccine to be successful, it needs to not only be proven as safe and efficacious, but also widely accepted.

It is estimated that a novel COVID-19 vaccine will need to be accepted by at least 55% of the population to provide herd immunity (Kwok 2020; Sanche 2020). Reaching these required vaccination levels should not be assumed given well-documented evidence of vaccine hesitancy across the world (de Figueiredo, 2020), which is often fuelled by online and offline misinformation surrounding the importance, safety, or effectiveness of vaccines (Bellaby 2003; Burki 2019; Lo 2017). As COVID-19 vaccine trials continue, there have been widely circulating (false) stories about the pandemic, including in the UK and US, such as that 5G mobile networks are linked with the virus, that vaccine trialists have died after taking a candidate COVID-19 vaccine, and that the pandemic is a conspiracy or a bioweapon (BMJ 2020; Geldsetzer 2020; Pennycook 2020). Such information builds on pre-existing fears over a new vaccine, seeding doubt and cynicism over a novel vaccine and threatens to limit public uptake of a COVD-19 vaccine.

While large-scale vaccine rejection threatens herd immunity goals, large-scale acceptance with local vaccine rejection can also have major consequences for community (herd) immunity as clustering of non-vaccinators can disproportionately increase the needed percentage of vaccination coverage to achieve herd immunity in adjacent geographical regions and encourage epidemic spread (Salathé 2008). Estimates of acceptance of a COVID-19 vaccine in June 2020 suggest that 38% of the public surveyed in the UK would “definitely” accept a COVID-19, while just 34.2% would in the US (a further 31% and 25% are, respectively, unsure about vaccinating) (McAndrew & Allington 2020). Worryingly, more recent polling in the US (September 2020) has shown significant falls in willingness to accept a COVID-19 for both males and females, all age groups, and all ethnicities (Pew Research 2020), likely due to the heavy politicisation of a COVID-19 vaccine in the run up to the 2020 Presidential Election (COCONEL 2020; Galvão 2020). The public’s willingness to accept a vaccine is therefore not static: it is highly responsive to current information and sentiment around a COVID-19 vaccine, as well as the state of the epidemic and perceived risk of contracting the disease. Under these current plausible COVID-19 vaccine acceptance rates, possible levels of existing protective immunity—though it is unclear whether post-infection immunity confers long-term immunity (Altmann 2020)—and the rapidly evolving nature of misinformation surrounding the pandemic (Pennycook 2020, WHO 2020), it is unclear whether we will reach vaccination levels required for herd immunity.

Although studies have examined the effect of COVID-19 misinformation on public pandemic perceptions (Geldsetzer 2020; Islam 2020; Kim 2020) and the tendency of certain socio-political groups to believe misinformation (Kreps 2020; Murphy 2020), we lack a quantitative understanding of the link between exposure to misinformation surrounding COVID-19 and intent to receive a future vaccination. As a viable vaccine comes closer to reality, it is essential to understand this link, how it differentially impacts socio-demographic groups, and whether groups at high risk of developing severe complications from COVID-19 are vulnerable to misinformation.

To fill this gap, we developed a COVID-19 questionnaire to measure vaccine intent pre- and post-exposure to online sources of recent misinformation relating to COVID-19 and vaccines. This questionnaire was used to survey 8,001 respondents across the UK and US via nationally representative sampling. 3,000 respondents in each country were exposed to misinformation (see Figure 1), while the remaining 1,000 were shown information about a COVID-19 vaccine that was factual to serve as a randomised control (see Figure 2 and Methods). A large suite of complementary data were collected for each individual including socio-econo-demographic status (age, gender, highest education level, employment type, religious affiliation, ethnicity, income level), sources of trust for information about COVID-19, political affiliation, social media usage, and reasons for being unsure about taking a COVID-19 vaccine (see Table 1 and Questionnaire, Appendix E).

**Table 1.**
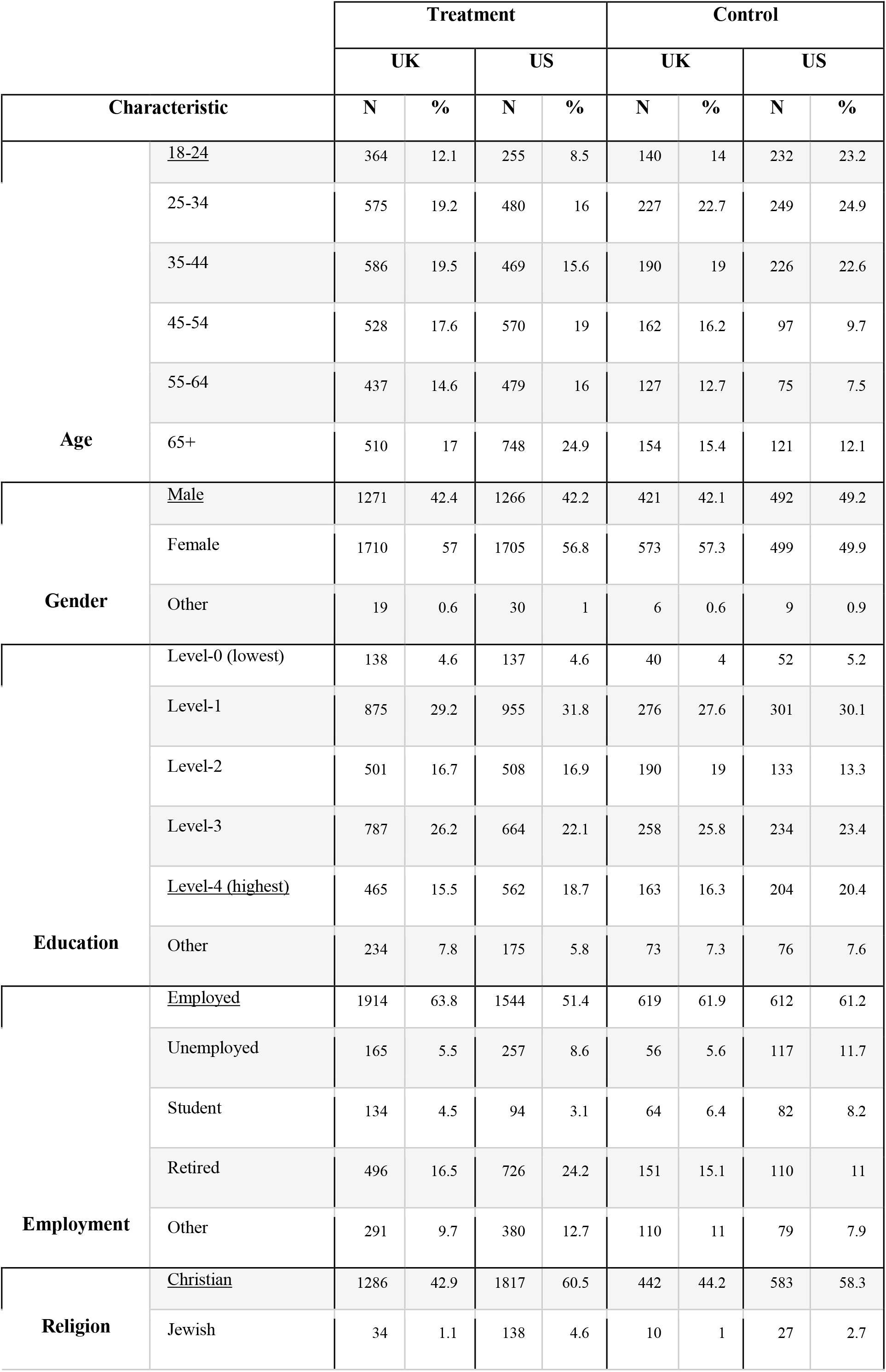
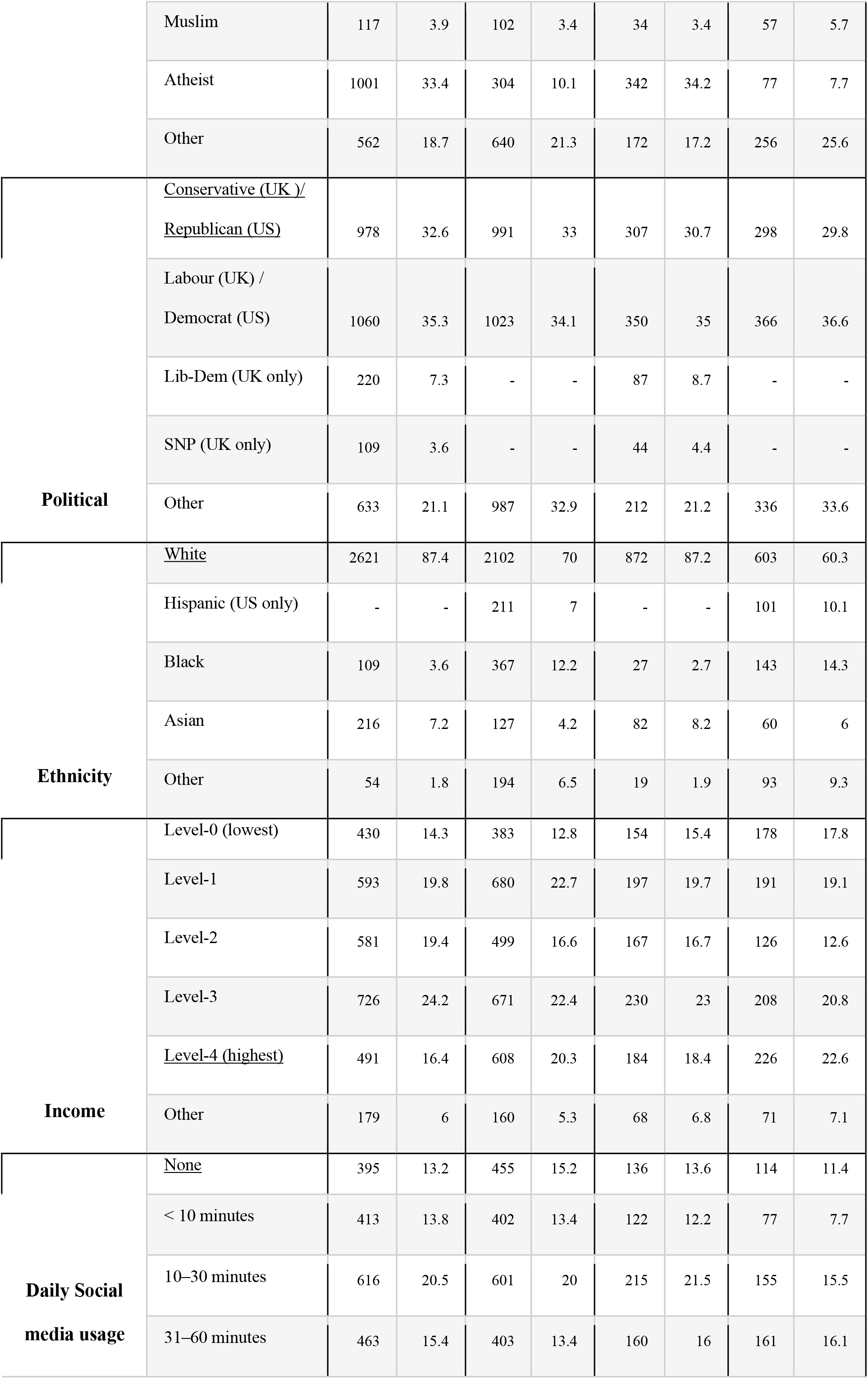
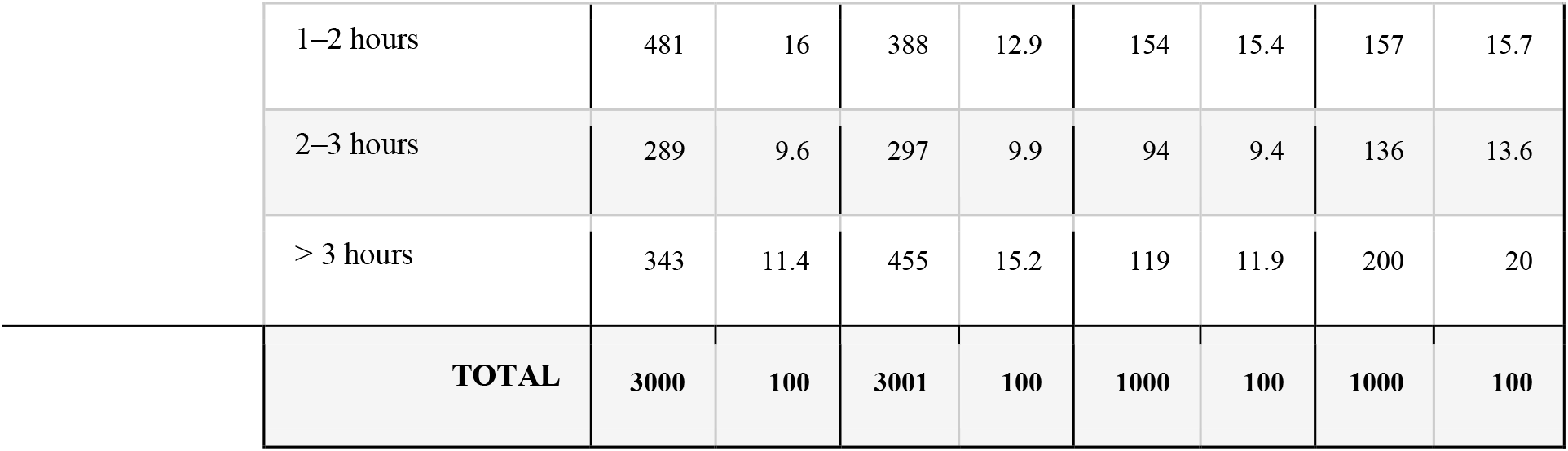
Socio-economic and characteristics of respondents. The reference (baseline) category used in the statistical modelling (see Methods) is underlined. The baseline category is: Male, 18-24, highest education, employed, Christian, White, Conservative (UK)/Republican (US), highest income, and no social media usage. Some socio-demographics have been recoded (see Appendix A for more details on variable recoding, and Appendix E for the original questionnaire).

|  |  | Treatment |  |  |  | Control |  |  |  |
| --- | --- | --- | --- | --- | --- | --- | --- | --- | --- |
|  |  | UK |  | US |  | UK |  | US |  |
| Characteristic |  | N | % | N | % | N | % | N | % |
| Age | <u>18-24</u> | 364 | 12.1 | 255 | 8.5 | 140 | 14 | 232 | 23.2 |
|  | 25-34 | 575 | 19.2 | 480 | 16 | 227 | 22.7 | 249 | 24.9 |
|  | 35-44 | 586 | 19.5 | 469 | 15.6 | 190 | 19 | 226 | 22.6 |
|  | 45-54 | 528 | 17.6 | 570 | 19 | 162 | 16.2 | 97 | 9.7 |
|  | 55-64 | 437 | 14.6 | 479 | 16 | 127 | 12.7 | 75 | 7.5 |
|  | 65+ | 510 | 17 | 748 | 24.9 | 154 | 15.4 | 121 | 12.1 |
| Gender | <u>Male</u> | 1271 | 42.4 | 1266 | 42.2 | 421 | 42.1 | 492 | 49.2 |
|  | Female | 1710 | 57 | 1705 | 56.8 | 573 | 57.3 | 499 | 49.9 |
|  | Other | 19 | 0.6 | 30 | 1 | 6 | 0.6 | 9 | 0.9 |
| Education | Level-0 (lowest) | 138 | 4.6 | 137 | 4.6 | 40 | 4 | 52 | 5.2 |
|  | Level-1 | 875 | 29.2 | 955 | 31.8 | 276 | 27.6 | 301 | 30.1 |
|  | Level-2 | 501 | 16.7 | 508 | 16.9 | 190 | 19 | 133 | 13.3 |
|  | Level-3 | 787 | 26.2 | 664 | 22.1 | 258 | 25.8 | 234 | 23.4 |
|  | <u>Level-4 (highest)</u> | 465 | 15.5 | 562 | 18.7 | 163 | 16.3 | 204 | 20.4 |
|  | Other | 234 | 7.8 | 175 | 5.8 | 73 | 7.3 | 76 | 7.6 |
| Employment | <u>Employed</u> | 1914 | 63.8 | 1544 | 51.4 | 619 | 61.9 | 612 | 61.2 |
|  | Unemployed | 165 | 5.5 | 257 | 8.6 | 56 | 5.6 | 117 | 11.7 |
|  | Student | 134 | 4.5 | 94 | 3.1 | 64 | 6.4 | 82 | 8.2 |
|  | Retired | 496 | 16.5 | 726 | 24.2 | 151 | 15.1 | 110 | 11 |
|  | Other | 291 | 9.7 | 380 | 12.7 | 110 | 11 | 79 | 7.9 |
| Religion | <u>Christian</u> | 1286 | 42.9 | 1817 | 60.5 | 442 | 44.2 | 583 | 58.3 |
|  | Jewish | 34 | 1.1 | 138 | 4.6 | 10 | 1 | 27 | 2.7 |
|  | Muslim | 117 | 3.9 | 102 | 3.4 | 34 | 3.4 | 57 | 5.7 |
|  | Atheist | 1001 | 33.4 | 304 | 10.1 | 342 | 34.2 | 77 | 7.7 |
|  | Other | 562 | 18.7 | 640 | 21.3 | 172 | 17.2 | 256 | 25.6 |
| **Political** | <u>Conservative (UK )/</u><br><u>Republican (US)</u> | 978 | 32.6 | 991 | 33 | 307 | 30.7 | 298 | 29.8 |
|  | Labour (UK) /<br>Democrat (US) | 1060 | 35.3 | 1023 | 34.1 | 350 | 35 | 366 | 36.6 |
|  | Lib-Dem (UK only) | 220 | 7.3 | - | - | 87 | 8.7 | - | - |
|  | SNP (UK only) | 109 | 3.6 | - | - | 44 | 4.4 | - | - |
|  | Other | 633 | 21.1 | 987 | 32.9 | 212 | 21.2 | 336 | 33.6 |
| **Ethnicity** | <u>White</u> | 2621 | 87.4 | 2102 | 70 | 872 | 87.2 | 603 | 60.3 |
|  | Hispanic (US only) | - | - | 211 | 7 | - | - | 101 | 10.1 |
|  | Black | 109 | 3.6 | 367 | 12.2 | 27 | 2.7 | 143 | 14.3 |
|  | Asian | 216 | 7.2 | 127 | 4.2 | 82 | 8.2 | 60 | 6 |
|  | Other | 54 | 1.8 | 194 | 6.5 | 19 | 1.9 | 93 | 9.3 |
| **Income** | Level-0 (lowest) | 430 | 14.3 | 383 | 12.8 | 154 | 15.4 | 178 | 17.8 |
|  | Level-1 | 593 | 19.8 | 680 | 22.7 | 197 | 19.7 | 191 | 19.1 |
|  | Level-2 | 581 | 19.4 | 499 | 16.6 | 167 | 16.7 | 126 | 12.6 |
|  | Level-3 | 726 | 24.2 | 671 | 22.4 | 230 | 23 | 208 | 20.8 |
|  | <u>Level-4 (highest)</u> | 491 | 16.4 | 608 | 20.3 | 184 | 18.4 | 226 | 22.6 |
|  | Other | 179 | 6 | 160 | 5.3 | 68 | 6.8 | 71 | 7.1 |
| **Daily Social**<br>**media usage** | <u>None</u> | 395 | 13.2 | 455 | 15.2 | 136 | 13.6 | 114 | 11.4 |
|  | < 10 minutes | 413 | 13.8 | 402 | 13.4 | 122 | 12.2 | 77 | 7.7 |
|  | 10–30 minutes | 616 | 20.5 | 601 | 20 | 215 | 21.5 | 155 | 15.5 |
|  | 31–60 minutes | 463 | 15.4 | 403 | 13.4 | 160 | 16 | 161 | 16.1 |

**Table 1 Socio-economic and characteristics of respondents.**
|  |  |  |  |  |  |  |  |  |
| --- | --- | --- | --- | --- | --- | --- | --- | --- |
| 1–2 hours | 481 | 16 | 388 | 12.9 | 154 | 15.4 | 157 | 15.7 |
| 2–3 hours | 289 | 9.6 | 297 | 9.9 | 94 | 9.4 | 136 | 13.6 |
| > 3 hours | 343 | 11.4 | 455 | 15.2 | 119 | 11.9 | 200 | 20 |
| **TOTAL** | **3000** | **100** | **3001** | **100** | **1000** | **100** | **1000** | **100** |

**Figure 1.**
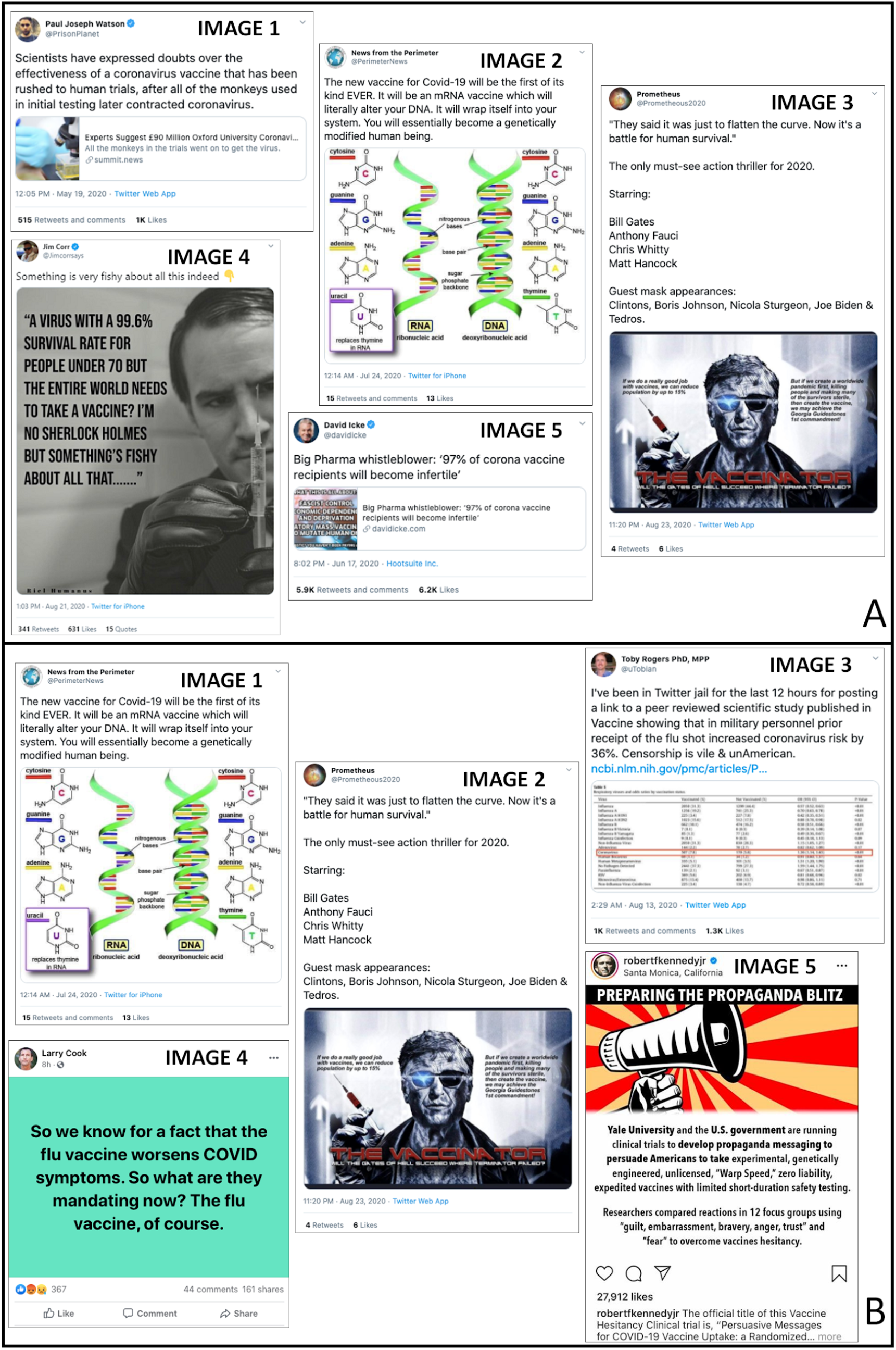
Widely circulating misinformation on social media surrounding a COVID-19 vaccine between June and August 2020. For each of the UK and US, five images were selected (see Methods) to expose to respondents. These “treatment” image sets were shown to 3,000 respondents in the UK (A) and the US (B). For reference, each image is numbered. URLs for all images are provided in Appendix B.

**Figure 2.**
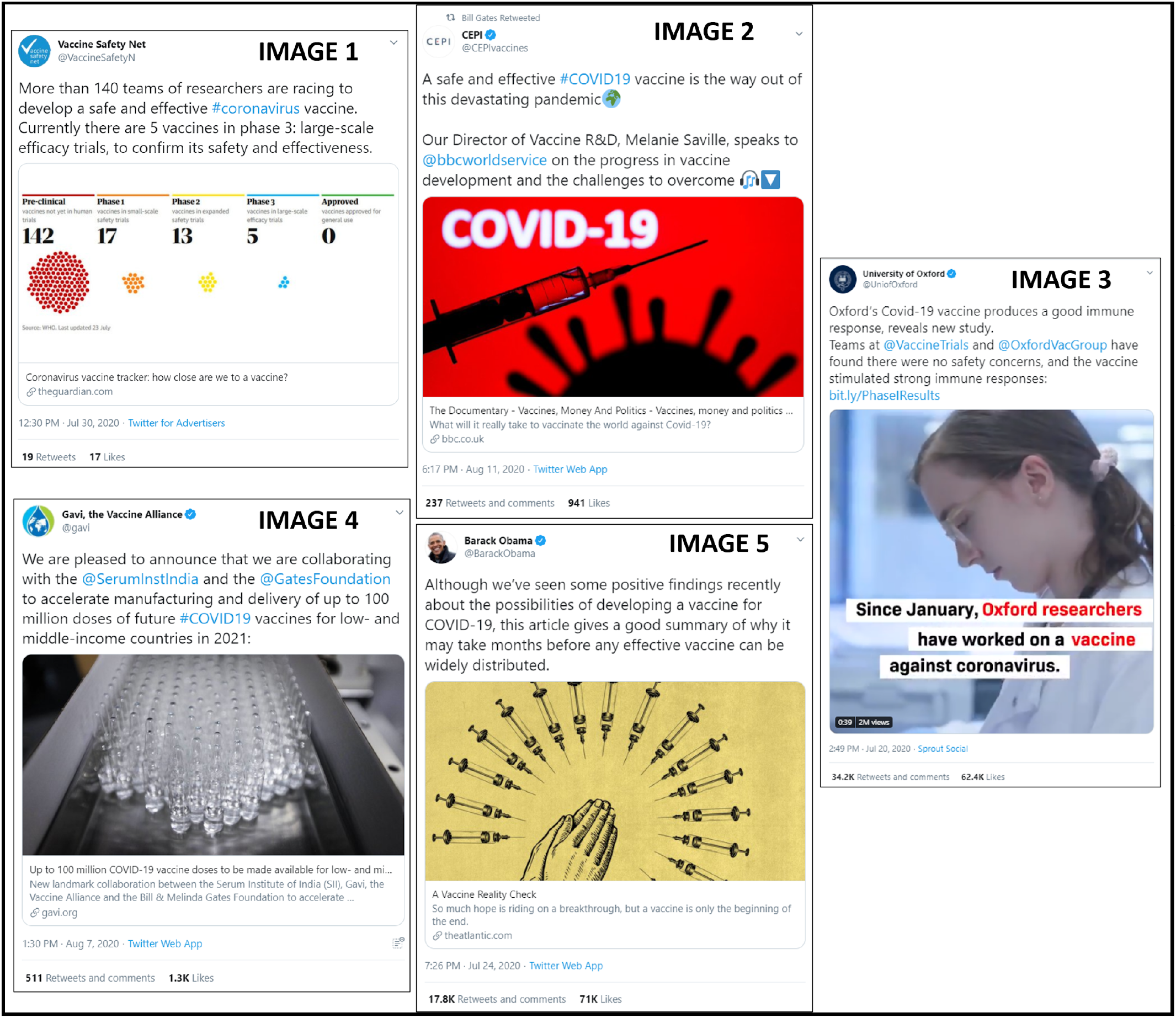
Widely circulating factual information on social media surrounding a COVID-19 vaccine between June and August 2020. The same five images were selected (see Methods) for exposure to respondents in the UK and US. These “control” image sets were shown to 1,000 respondents in the UK and the US. URLs for all images are provided in Appendix B.

To assist policymakers and stakeholders in the design of communication and messaging campaigns, we calculate the determinants of COVID-19 vaccine intent both pre-exposure *and* post-exposure, allowing us to establish the groups who are already less likely to vaccinate *and* those who are susceptible to COVID-19 vaccine misinformation.

We also consider how intention to vaccinate is driven by motivation to protect oneself, family, friends, and at-risk groups and how this changes after exposure to misinformation. By exploring vaccination intent to protect others, we are able to quantify how misinformation may impact altruistic vaccination behaviour: particularly important in the UK and US where altruistic messaging prompts have been a feature of COVID-19 public health messaging campaigns (NHSa 2020; NHSb 2020; CDCa 2020; CDCb 2020). Moreover, an assessment of the impact of different types of misinformation presented to respondents yields an understanding of the semantic and stylistic content of misinformation that has the largest impact in lowering intent to vaccinate in this study.

Our findings are interpreted in light of vaccination levels required for herd immunity and we discuss messaging strategies that may help mitigate or counter the impact of online vaccine misinformation. Throughout this study, misinformation refers to information that is inadvertently false, but no harm is meant in sharing it and that “[is] considered incorrect based on the best available evidence from relevant experts at the time” (Wardle 2017, Vraga 2020).

## Results

The questionnaire was fielded between 7 and 14 September, 2020 to a total of 8,001 respondents. All respondents were recruited via an online panel by ORB (Gallup) International (www.orb-international.com). Respondent quotas were set according to national demographic distributions for gender, age, and sub-national region (state in the US and first level of nomenclature of territorial units in the UK (ONS 2020). A total of 4,001 participants were surveyed in the UK and 4,000 in the US, and all respondents were aged 18 or over. Of these 8,001 respondents, 3,001 (3,000) respondents were exposed to misinformation relating to COVID-19 and vaccines (treatment group) in the UK (US) and 1,000 were exposed to factual COVID-19 vaccine information (control group).

All information (misinformation and factual) was identified using Meltwater^®^ (www.meltwater.com) via a Boolean search string eliciting (mis)information around a COVID-19 vaccine. A systematic selection approach was used to identify the COVID-19 vaccine information on social media with high circulation and engagement between 1 June, 2020, and 30 August, 2020 (see Methods). A final set of five pieces of misinformation comprising non-overlapping messaging and themes were selected to represent the diverse messaging found in COVID-19 vaccine misinformation (such as information questioning the importance or safety of a vaccine, see Figure 1). As misinformation can be highly country and context-dependent, it was decided to expose UK and US respondents to different sets of misinformation to reflect the different audiences targeted by the sources of misinformation, while factual information was the same for both. Each piece of (mis)information was shown on a separate page to facilitate image comprehension.

Respondents were asked about their intent to receive a COVID-19 vaccine to protect themselves and, to explore altruistic behaviours, to protect others: “If a new coronavirus (COVID-19) vaccine became available, would you accept the vaccine for yourself?” (SELF) and “if a new coronavirus (COVID-19) vaccine became available, would you accept the vaccine if it meant protecting friends, family, or at-risk groups?” (OTHER). Responses were collected on a four-point scale: “Yes, definitely,” “unsure, but leaning towards yes,” “unsure, but leaning towards no,” and “no, definitely not.” This scale is chosen to remove subjective ambiguity involved with Likert scales and to allow respondents to detail explicitly their intent, thereby allowing a more meaningful interpretation of results. Both questions (SELF and OTHER) were asked to respondents before and after exposure to either the treatment or control information set.

### The impact of misinformation on vaccination intent

We first investigated individuals’ intent to vaccinate before and after information exposure. Before any exposure, 54.0% (95% percentile interval [PI], 52.3 to 55.9) of respondents in the UK and 41.2% (39.0, 43.0) in the US report that they will “definitely” accept a COVID-19 vaccine to protect themselves (Table 2). Higher intent to accept a COVID-19 in the UK than the US has been recently reported (McAndrew 2020).

**Table 2.**
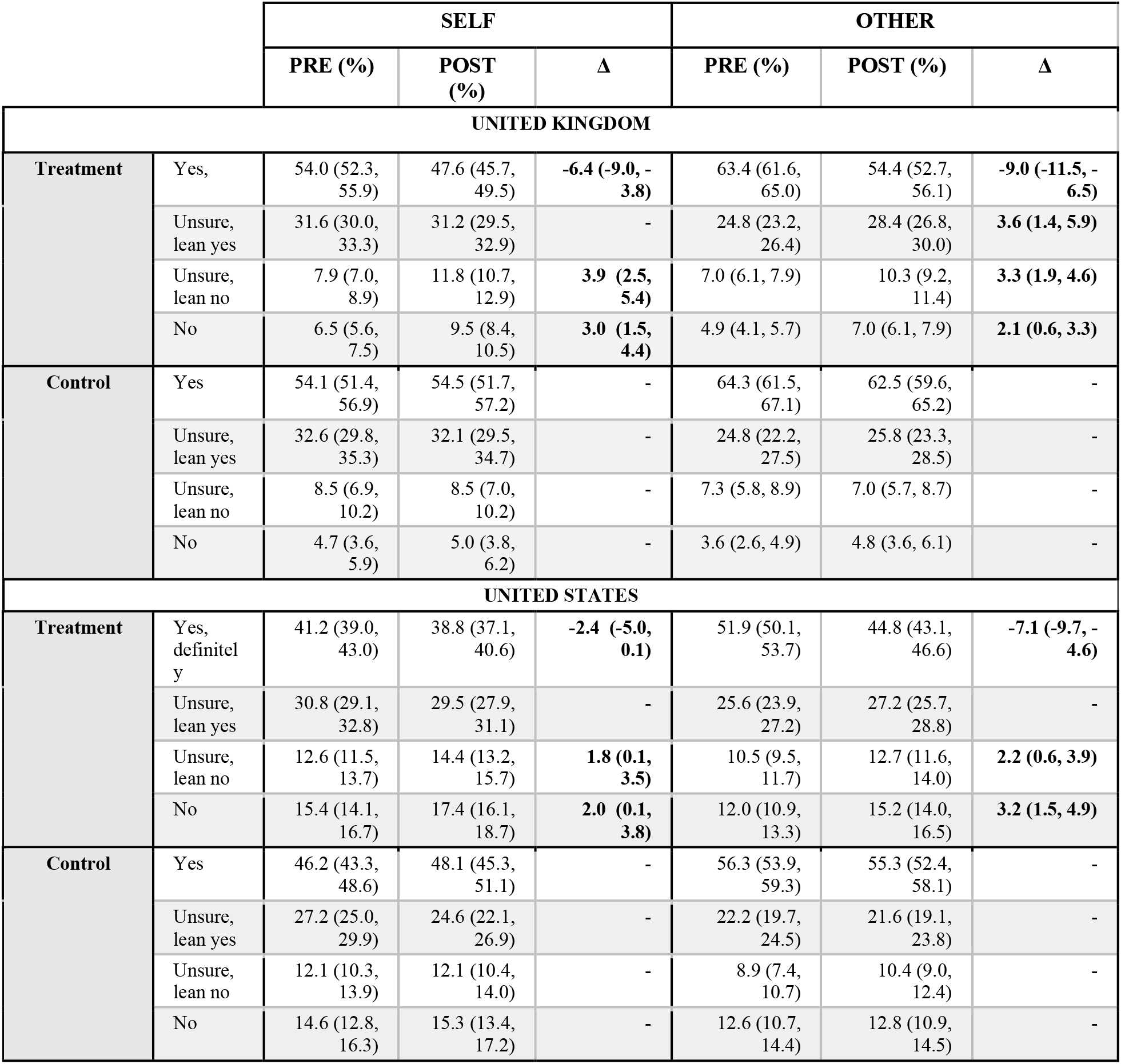
Exposure to misinformation reduces intent to vaccinate. Percentage of respondents in the UK and US reporting that they would definitely accept, definitely not accept, or were unsure about accepting a COVID-19 vaccine for themselves (SELF) or others (OTHER) pre- and post-exposure to factually incorrect information about COVID-19 (treatment) or factually correct information (control). Numbers in parentheses denote the upper and lower 95% percentile interval. Only significant pre- and post-exposure changes (Δ) are shown.

Exposure to misinformation induces significant decreases in the number of respondents reporting that they would “definitely” accept a vaccine: a 6.4 (3.8, 9.0) percentage point (pp) fall in the UK and a 2.4pp (0.1, 5.0) fall in the US. There are corresponding increases in the percentages of respondents either now unsure whether to vaccinate or who would now definitely not vaccinate (Table 2). We find that factual information induces no “significant” shifts (95% PI of difference distribution includes zero) in the proportion of respondents falling into each response category before and after exposure (Table 2).

We find two results that may be of particular interest to policymakers harnessing altruistic messaging devices to boost public compliance with recommended interventions. Firstly, more respondents in both countries would accept a vaccine if it meant protecting family, friends, or at-risk groups (than if the vaccine was for themselves): 63.4% (61.6, 65.0) of respondents in the UK and 51.9% (50.1, 53.7) in the US say that they would “definitely” get vaccinated to protect others (Table 2). Secondly, misinformation induces a disproportionately larger percentage point fall in vaccination intent to protect others than to protect oneself: a 9.0pp fall in the UK (a 14.2% percentage drop compared to 11.9% for the drop associated with intent to vaccinate themselves) and a 7.1pp fall in the US (13.7% versus 6.0%). These results suggest that misinformation may have a damaging effect on messaging drives that leverage individual altruistic behaviours.

While the above analysis reveals exposure to misinformation impacts overall vaccine intent, we cannot casually attribute these changes to the misinformation shown to respondents without accounting for the control group. We therefore estimate the risk difference of transitioning from one vaccine acceptance category to another upon exposure to misinformation, relative to exposure to factual information—see Figures S1, S2 and Tables S1, S2 in Appendix D. These risk differences (as defined in Equation 2 of Appendix C) reveal that there is still an overall transition of people from categories of higher to lower vaccine acceptance when controlling for exposure to factual information.

### Determinants of COVID-19 vaccination intent

Bayesian ordinal logistic regressions (see Methods) were used to find the factors associated with rejecting a COVID-19 vaccine and the factors associated with a susceptibility to misinformation—that is, a tendency for individuals to *lower* their vaccination intent after seeing misinformation. There are some key similarities and differences between the determinants in the UK and the US of relevance to country-specific public health policy, which we outline here. Effects are reported as odds ratios (OR), where odds ratios larger than 1 represent a factor that is associated with increased chance of rejecting the vaccine compared to the baseline group (Male, 18-24, highest education, employed, Christian, White, Conservative (UK)/Republican (US), highest income, and no social media usage).

In both countries, females are more likely than males to refuse a COVID-19 vaccine, with a larger effect-size in the US (odds ratio 2.02, 95% percentage interval (PI): 1.78 to 2.29) than the UK (OR 1.44 [1.25, 1.63]). (All odds ratios and credible intervals are shown in Figure 3 for UK and US pre-exposure determinants (A, C) and determinants of susceptibility (B, D).) In both countries, individuals with highest educational attainment below postgraduate degrees, low income groups, and non-whites are more likely to reject a COVID-19 vaccine. Trends in uptake intent determinants are found to differ along age, employment status, political affiliation, and social media use. Most notable among these differences are that 55-64 year-olds and over 65s are less likely to reject a COVID-19 vaccine than 18-24 year-olds in the UK, whereas all age groups except over 65s are more likely to reject a vaccine than 18-24 year-olds in the US. In the US, Democrats are less likely to reject the vaccine than Republicans (OR 0.66 [0.56, 0.76]) whereas those who do not affiliate with any of the four major parties in the UK (Conservative, Labour, Lib Dems, SNP) are far more likely to refuse a vaccine. These results mirror recent surveys in Ireland, where COVID-19 vaccination intent is associated with non-mainstream political affiliation (Murphy et al. 2020), and in the US, where males and those with degrees were more likely to accept the vaccine (Malik 2020), and where Democrat voters are “more likely to correctly identify a Covid-related headline as true or false than were independents or Republicans” (Kreps 2020). Individuals in the US with no social media use are less likely to accept a vaccine than individuals who use social media. In the US, individuals with low social media usage (and high legacy media usage) have higher vaccine intent than those with low consumption of both (McAndrew 2020). (Full regression results are provided in Appendix D Table S2.)

**Figure 3.**
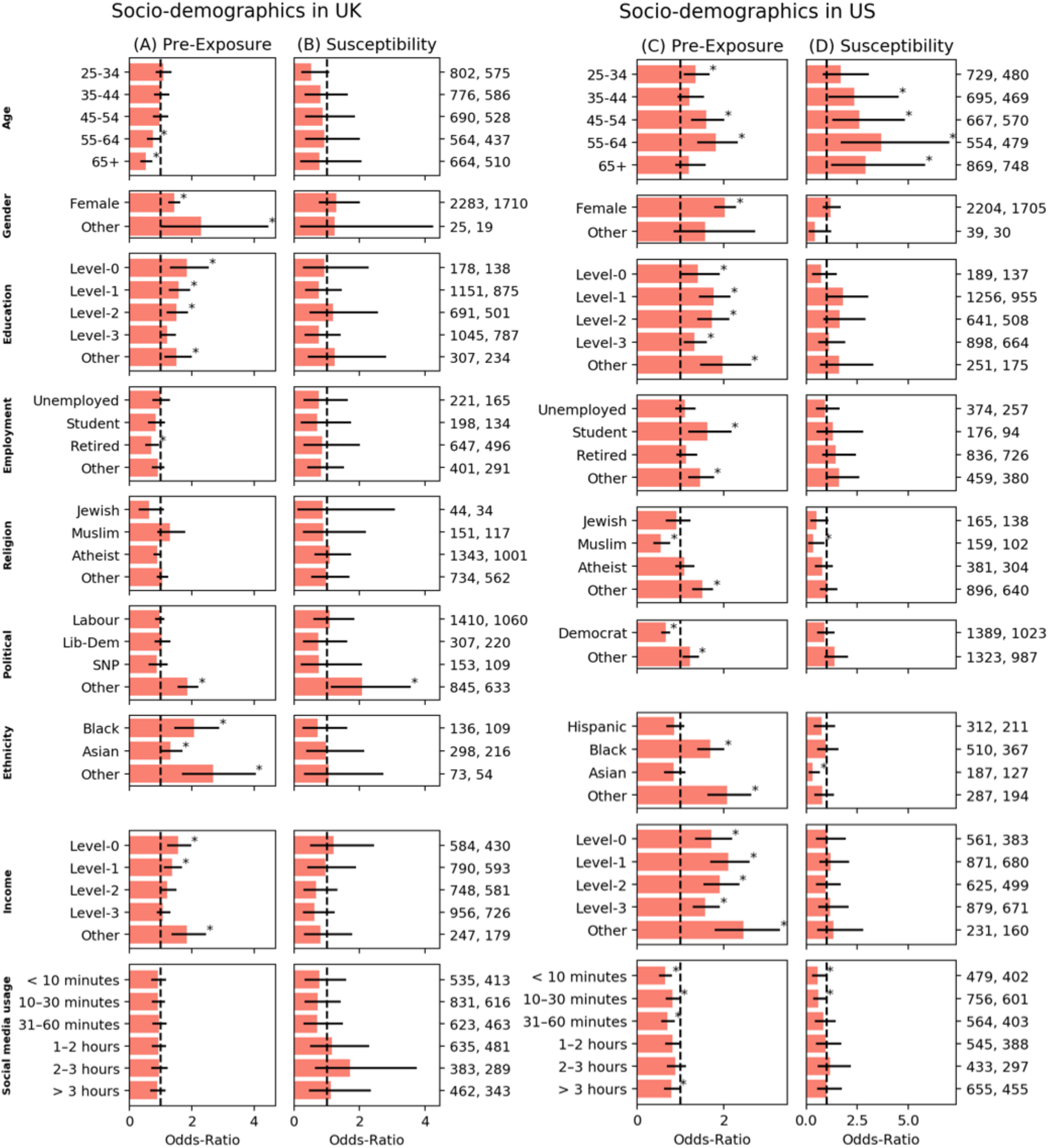
Determinants of vaccination intent before exposure and of susceptibility to misinformation. Contribution of socio-demographic and social-media-use characteristics to pre-exposure vaccine intent (columns A, C) and susceptibility to misinformation exposure (columns B, D) for the UK (Left) and US (Right). Red bars depict odds-ratios (OR) of being more reluctant to vaccinate or more susceptible to misinformation relative to the reference category of Male, 18-24, highest education, employed, Christian, White, Conservative, highest income, and no social media usage. Odds ratios above 1 indicate the group is more likely to reject a COVID-19 vaccine than the reference group, and more likely to be susceptible to misinformation relating to COVID-19 and vaccines. Black bars indicate 95% percentile intervals and starred (*) bars show effects whose 95% PI excludes zero (which we term “significant”). Numbers on the right indicate sample sizes of the corresponding demographic. See Table S3 in Appendix D for more details.

Many groups in the US appear to be vulnerable to COVID-19 vaccine misinformation. We find that respondents aged over 35 are significantly more likely to reject a vaccine after exposure to misinformation than they were before (OR 2.35 [1.08, 4.54] for 35-44 years, OR 2.62 [1.28, 4.84] for 45-54, OR 3.68 [1.70, 7.03] for 55-64 and OR 2.90 [1.23, 5.85] for 65+). In addition, Asians (OR 0.31 [0.11, 0.68]) and Muslims (OR 0.34 [0.10, 0.89]) are less susceptible to misinformation than Whites and Christians (respectively). Interestingly in the US, those who use up to 30 minutes of social media are less susceptible than non-users of social media (OR 0.57 [0.29, 0.99] for less than 10 minutes of social media use per day and OR 0.60 [0.34, 0.99] for 10-30 minutes per day).

In the UK, the impact of misinformation on intent appears to have no differential effect across socio-econo-demographic groups, but those not supporting one of the four main political parties are more susceptible to misinformation 2.08 [1.13, 3.57] (compared to the Conservatives).

### Reasons for COVID-19 vaccine hesitancy

Respondents who would not “definitely” take a COVID-19 vaccine were asked for their reasons, which were included as explanatory factors in the ordinal logistic regressions used previously, while controlling for socio-demographic factors (see Methods). In this section the log odds ratio (log OR) is used: log ORs larger than 0 represent a factor that is associated with increased chance of rejecting the vaccine, and of being more susceptible to misinformation, compared to the baseline group (same as previously stated with no social media baseline).

In the UK and US, vaccine rejection is associated with a belief that COVID-19 does not pose a risk or that they will not be ill if they contracted the disease (Figure 4A, B). Such barriers fall within the “complacency” vaccine hesitancy under WHO’s Strategic Advisory Group of Experts (SAGE) on immunisation’s “3C” hesitancy model (WHO 2014). Respondents who claimed that they wanted to “wait until others” received the vaccine, were more likely to lean towards vaccinating (“unsure, but leaning towards yes”) than respondents who said they were “unsure, but leaning towards no” or would “definitely not” vaccinate (log odds ratio, - 0.41 [-0.67, −0.17] for the UK and log OR −0.50 [−0.69, −0.31] for the US).

**Figure 4.**
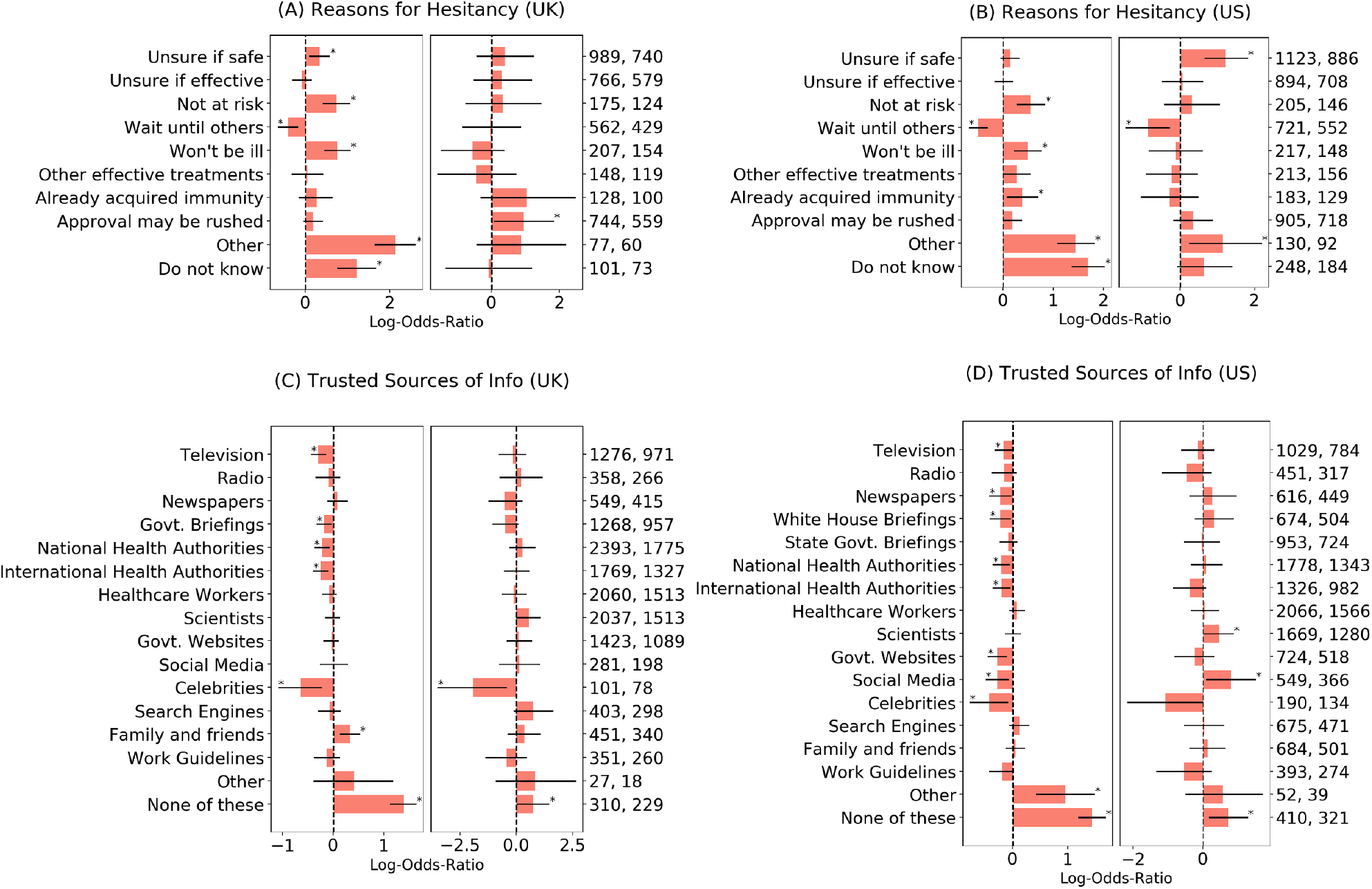
Concerns over vaccine safety and a mistrust in mainstream sources of information are drivers COVID-19 vaccine rejection. Contribution of reasons that respondents provide for not being “definitely” sure of taking a COVID-19 vaccine (A, B) and contribution of sources of information that people trust (C, D), to the Pre-Exposure vaccine hesitancy (left of every sub-figure) and Susceptibility to vaccine misinformation (right of every sub-figure) as measured by drop in vaccine intent—after controlling for socio-demographics. Values depict odds-ratios (OR) of being more hesitant (for pre-exposure) or more susceptible (for susceptibility)—as measured relative to when the reason was not indicated for hesitancy, or when the source was not indicated as being trusted. **OR>1 indicates the group is more likely to not accept a COVID-19 vaccine if they indicated that reason for hesitancy (A, B) or that source of information as trustworthy (C, D)**. Bars indicate 95% percentile intervals and * indicate “statistical significance” based on them. Numbers on the right indicate sample sizes that had indicated the corresponding reason/source, within both the total (N=4000 for UK and N=4001 for US) and treatment respondent set (N=3000 for UK and N=3001 for US). Since reasons for hesitancy were only asked to those who did not choose “Yes, definitely” when asked if they would vaccinate to protect themselves, that analysis is conditioned to those who did not indicate “Yes, definitely” for the SELF question.

Respondents who report that “vaccine approval may be rushed” tent to be significantly more vulnerable to misinformation in the UK (log OR 0.93 [0.07, 1.83]) (Figure 4A), while, in the US, respondents who are unsure whether the vaccine is safe are more susceptible to misinformation (log OR 1.22 [0.65, 1.82]) (Figure 5B). These two responses are both related to explicit “confidence” barriers (WHO 2014).

**Figure 5.**
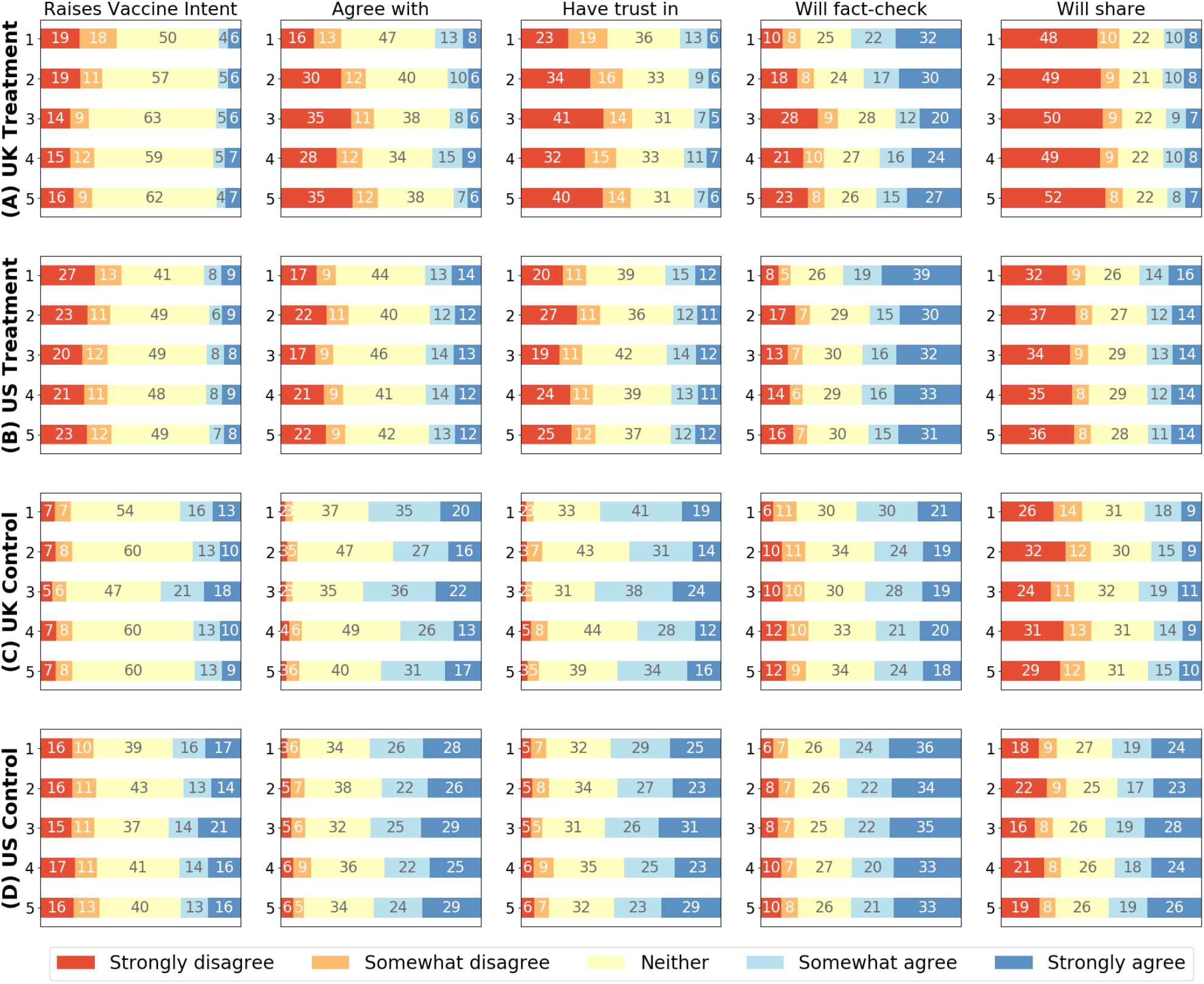
Breakdown of views to information displayed to respondents. The percentage of respondents providing a given response to each follow-up question to explore their perceptions of each image. Respondents were asked whether each image raises their vaccine intent (column 1); contains information they agree with (column 2); contains information they find trustworthy (column 3); is likely to be fact-checked by them (column 4); and is something they will likely share with others (column 5). Rows represent images shown to (A) UK treatments, (B) US treatments, (C) UK controls and (D) US controls. Note that those responding with “Do not know” were grouped with those saying “neither/nor”. 95% percentile intervals have been excluded here for clarity. Note that the response scale has been inverted for column 1 for direct comparison across all questions (see Section 4 of Appendix E for the relevant questionnaire subsection).

### Sources of information that are trusted

Respondents across both countries who trust television news, government briefings, health authorities, and (perhaps surprisingly) celebrities tend to have higher pre-exposure inclination to vaccinate. Respondents who indicate that they trust family and friends have lower pre-exposure intent than those who do not trust family or friends in the UK (log OR 0.33 [0.12, 0.53]). Respondents in both countries who report trusting no sources (“None of the above”, see Table S5, Appendix D) have lower pre-exposure vaccination intent (see Figure 4C, D). Those who do not trust any typical sources remain significantly more susceptible to misinformation exposure in both the UK and US (Figure 4C and D), suggesting that a lack of trust in mainstream authorities is indicative of being influenced by misinformation exposure. Trust in social media and scientists also contributes to susceptibility for the US, but not in the UK.

### Qualitative assessment of the appeal of scientific misinformation

After exposure to misinformation or factual information, respondents were asked to report whether, for each image: it raised their vaccination intent; they agreed with the information presented; they found the information to be trustworthy; they were likely to fact-check; and they were likely to share the image with friends or followers.

Across both countries, sizeable proportions (between 13% and 27% of those who would not definitely accept the vaccine) either agreed with the misinformation or found it trustworthy, though the majority of respondents did not agree and did not find it trustworthy (Figure 5A, B). To investigate the impact that each individual image had on vaccine intent, weights were assigned to each image while regressing image characteristics—self-reported by respondents—against changes in vaccine intent upon exposure, thus quantifying the contribution of each piece of (mis)information to the change (see Methods). This analysis revealed that the images with the largest contribution to a loss in vaccination intent in the UK were Image 1 (“scientists have expressed doubts […] over the coronavirus vaccine […] after all of the monkeys used in initial testing contracted coronavirus”) and Image 2, which claimed the new COVID-19 vaccine will “literally alter your DNA” (Appendix D, Table S6 and S7). This latter image was the most impactful in lowering vaccination intent in the US, followed by Image 5 “Yale University and the U.S. government are running clinical trials to develop propaganda messaging to persuade Americans to take […] vaccines […]” (Appendix D, Table S6 and S7). In the control set—which was shown to respondents in both countries— the image that contributed the *least* to fall in vaccine intent was Image 3 (Figure 2), in which the University of Oxford announced that their vaccine “produces a good immune response” and that the “teams @VaccineTrials and @OxfordVacGroup have found there were no safety concerns”. While other images arguably used some scientific messaging (such as Image 5 in Figure 1A, “Big Pharma whistleblower: ‘97% of corona vaccine recipients will become infertile’”), the images identified as having the most impact on lowering vaccination intent stated a direct link between the COVID-19 vaccine and adverse effects and cited articles and scientific imagery or links to articles purporting to be reputable to strengthen their claim. This contrasted to more memetic imaging (for example, “striking images with text superimposed on top” (Wardle 2017)) which was far less impactful (Images 3 and 4, Figure 1A and Images 2 and 4, Figure 1B).

The manner of information exposure in our study is a simplification of how people are naturally exposed to information online social media platforms. We asked respondents if they had encountered similar images to the ones they were exposed to on social media in the past one month, to explore the relationship between their vaccination intents and pre-study exposure to misinformation or factual information. We find evidence of misinformation and factual information “filter bubbles”, which is the phenomenon of people seeing more of the content that agrees with their prior beliefs (Pariser 2011, Bakshy 2015). In the UK, respondents who reported that they would definitely not vaccinate to protect themselves before exposure to misinformation were 10.6% (3.0, 18.1) more likely than those who would “definitely” get vaccinated to have seen similar misinformation. In both the UK and US, respondents in the control group who reported that they would “definitely not” vaccinate were significantly less likely to have seen similar factual information (see Appendix D, Table S8 and Figure S3).

## Discussion

Using individual-level survey data collected from nationally representative samples of 4,000 respondents in each of the UK and US, we reveal a number of key findings of importance to policymakers and stakeholders engaged in either public health communication or the design of vaccine-rollout programmes. We find that, as of September 2020, only 54.0% of the public in the UK and 41.2% in the US would “definitely” accept a COVID-19 vaccine to protect themselves. The main barriers to reporting certainty over vaccinating were concerns over vaccine safety or a belief that they would not be at risk of contracting COVID-19 or would not be ill if they did (that is, vaccine-importance related confidence barriers). Individuals who wanted to “wait until others” had been vaccinated were less likely to outright reject a vaccine.

Currently, these values are below those required to achieve the anticipated herd immunity levels, however, higher proportions of individuals in both countries would “definitely” vaccinate to protect family, friends, and at-risk groups, suggesting that effective altruistic messaging may be required to boost uptake. However, we have also shown that exposure to misinformation not only lowers intent to vaccinate, but that it disproportionately lowers intent to vaccinate to protect others, which could complicate messaging campaigns focusing on altruistic behaviours. Campaigns may also have to battle with misinformation purporting to be based in science or medicine, which appears to be particularly damaging on vaccination intentions.

These findings are, however, unlikely to be representative of the effect of misinformation on uptake rates in real world social-media settings. Individuals are unlikely to experience misinformation in the same manner as implemented in this survey, and there will be differences in the volume and rate of misinformation people will be exposed to, depending on their online social media preferences and demographics. A demographic re-weighting would be required to obtain more robust estimates of anticipated COVID-19 vaccine rejection at sub-national or national levels. Misinformation may have also already embedded itself in the public’s consciousness, and studies have shown that brief exposure to misinformation can embed itself into long-term memory (Zhu 2012). Policymakers may therefore find challenges ahead to “undo” the impact it may have already had and to clearly communicate messages surrounding the safety, effectiveness, and importance of the vaccine.

Willingness to accept a COVID-19 vaccine and susceptibility to misinformation is found to depend on a number of socio-economic factors (these findings are likely to be more robust since the survey ensures consistent misinformation exposure across different demographics). Females, ethnic minority groups, those without university degrees, and low-income groups were less willing to accept a vaccine in both the UK and US (with respect to the baseline, see Table 1), in alignment with recent studies (Sherman 2020; McAndrew & Allington 2020; Murphy 2020). Given that COVID-19 incidence and mortality rates are higher in some black and minority ethnic groups as well as lower-income groups in the UK and US (Aldridge 2020; Pan 2020; Patel 2020; Price-Haywood 2020), it is vital that vaccination rollout campaigns not only ensure sufficient access to these communities, but that confidence in the vaccine is built before rollout. The groups most susceptible to misinformation are older age groups in the US. Given that age is an important risk factor for COVID-19, an increase in misinformation targeted at these groups could prove detrimental to efforts in protecting those who are most at-risk. In the UK, older age groups appear to be more confident in the vaccine—consistent with previous studies into both COVID-19 perceptions (Murphy 2020) and longer-term general vaccine confidence trends in the UK (Larson 2018).

Respondents who did not report even a single source of trust were significantly more susceptible to misinformation than those who did, echoing research from the UK and Ireland which found that “those who were resistant to a COVID-19 vaccine were less likely to obtain information about the pandemic from traditional and authoritative sources and had similar levels of mistrust in these sources” (Murphy et al. 2020). In the UK, individuals who believed the vaccine may be rushed were the most negatively impacted by vaccine misinformation.

Although our study indicates the possible impact of COVID-19 misinformation campaigns on vaccination intent, this study does not replicate a real-world social media platform environment—where information exposure is a complex combination of what is shown to a person via the platform’s algorithms and what is shared by their friends or followers (Bakshy 2015). Online social network structures, governed by social homophily, can lead to selective exposure and creation of homogeneous “echo-chambers” (Del Vicario 2016) which may amplify (or dampen) the spread of misinformation among certain demographics. Indeed, we do find some evidence in this study of filter bubbles with regards to information on COVID-19 vaccine—those who would definitely not accept the vaccine had seen more misinformation (in the UK) and less factual information (in both the UK and US) recently online when compared to those who definitely would accept the vaccine. These network effects cannot be replicated via a questionnaire. Therefore, our estimates for the losses in vaccination intent due to misinformation must be placed in the context of this study: caution must be exercised in generalising these findings to a real-world setting, which may see greater or lesser drops in vaccination intent depending on the wider context of influencing factors. Moreover, we are limited in the type and volume of misinformation presented to respondents and there may exist other types of misinformation which may be far more impactful on vaccination intent. Addressing the spread of misinformation will likely be a major component of a successful COVID-19 vaccination campaign, especially as misinformation on social media has been shown to spread faster than factually correct information (Vosoughi 2018) and that, even after a brief exposure, misinformation can result in long-term attitudinal and behavioural shifts (Pluviano 2017; Zhu 2012) that pro-vaccination messaging may find hard to overcome (Pluviano 2017). With regards to COVID-19, misinformation has even been shown to lead to information avoidance and less systematic processing of COVID-19 information (Kim 2020), however, the amplification of “questionable” sources of COVID-19 misinformation is highly platform dependent, with some platforms amplifying questionable content less than reliable content (Cinelli 2020).

The analysis reveals that, in both the UK and US, fewer people would “definitely” take a vaccine than is required for herd immunity, and that misinformation could push these levels further away from herd immunity targets. This analysis provides a platform to help us test and understand how more effective public health communication strategies can be designed and on whom these strategies would have the most positive impact in countering COVID-19 vaccine misinformation.

## Methods

### Participants

A total of 8,001 participants were recruited via an online panel by ORB (Gallup) International (www.orb-international.com) and surveyed between 7 and 14 September, 2020 and who obtained informed consent was obtained from all participants. A total of 4,001 participants were surveyed in the UK and 4,000 in the US. Of these respondents, 3,001 (3,000) respondents were exposed to COVID-19 misinformation around vaccines (the **treatment** group) in the UK (US), while 1,000 in each country exposed to factual information relating to COVID-19 vaccines (the **control** group). Each group was shown a total of five pieces of information: all of these five were either misinformation or factual. Respondents were sampled to match proportions of national demographic breakdowns for gender, age, and sub-national region (state in the US and second administrative level in the UK (ONS 2020)). Survey weights were provided to account for mis-matches between expected national sex, age, and regional distributions and those obtained via online panels. Survey sizes for both the exposure and control exceed those typically conducted as national-level surveys (n=1000) and therefore summary variables contain at most a ∼3% error.

### Survey Design

Respondents are asked about their intent to vaccinate to protect themselves and (separately) their intent to vaccinate themselves to protect friends, family, and at-risk groups contracting COVID-19. To deduce changes in vaccination intent, respondents are asked for their intent to vaccinate both before and after exposure. The questions were as follows,

1. “if a new coronavirus (COVID-19) vaccine became available, would you accept the vaccine for yourself?” (**SELF**), and
2. if a new coronavirus (COVID-19) vaccine became available, would you accept the vaccine if it meant protecting friends, family, or at-risk groups?” (**OTHER**).

These two questions are answered on a four-level scale: “Yes, definitely”, “unsure, but leaning towards yes”, “unsure, but leaning towards no”, and “No, definitely not”. This scale is chosen to remove ambiguity involved with Likert scales and to provide a meaningful interpretation of results with regards to vaccination intent. If respondents do not assert that they would “yes, definitely” take a vaccine, then reasons for hesitancy are explored (see Figure 4A, B).

The information exposed to respondents are screenshots of social media posts, which contain a mixture of text and images (see Figures 1 and 2). A different set of five treatment images are shown to respondents in the UK and US to reflect country-specific differences in online social media sources and content, while the control images are a set of five images fixed for both countries (see *Methods: Selection of Intervention Images* for further information). For each exposure image, respondents are asked to rate the extent that: 1) they agree with the information displayed; 2) they are inclined to be vaccinated; 3) they believe the information to be trustworthy; 4) they will fact-check the information; and 5) they would share the image. After exposure, the respondents were also asked if they had seen “similar” content on social media in the last one month. (See Appendix E for the full questionnaire.)

Respondents’ socio-demographic information is collected, including gender, age, education, employment status, religious affiliation, ethnicity, income and political affiliation. To probe relationships with social media use, respondents are asked to self-report the amount of time spent daily on social media platforms (Ernala 2020). Raw counts for these characteristics are shown in Table 1 (see Appendix A for details on variable recoding). These characteristics are used to determine the groups most reluctant to take a COVID-19 vaccine and the groups most impacted by misinformation. In addition, respondents are asked to report the sources they trust for information surrounding COVID-19.

### Selection of Intervention Images

In order to elicit responses that can be most readily interpreted in light of the current state of online misinformation in both countries, the information shown to respondents should satisfy a number of criteria, it should: 1) be recent and relevant to a COVID-19 vaccine; 2) have a high engagement, either through user reach or other publicity, and thus represent information that respondents are not unlikely to be exposed to through social media use; 3) include posts shared by organisations or people with whom respondents are familiar (so that, for example, US/UK audiences are not shown information from people with whom they are unfamiliar); 4) form a distinct set, not replicating content or core messaging, allowing us to probe the most impactful types of misinformation.

To this end, we followed a principled approach to selecting five images, combining both quantitative and qualitative methods. The procedures for selecting treatment and control image sets are as follows:

(*Treatment Set*) We used a COVID-19 vaccine-specific Boolean search query—corona* OR coronavirus OR covid* OR “wuhan virus” OR wuhanvirus OR “chinese virus” OR “china virus” OR chinavirus OR “nCoV*” OR SARS-CoV*) AND vaccin* AND (Gates OR 5G OR microchip OR “New World Order” OR cabal OR globali*)—to extract COVID-19 vaccine related online images from 1 June, 2020, to 30 August, 2020 using Meltwater^®^ (www.meltwater.com), an online social media listening platform. This Boolean search term was based on previous research which used similar search terms which obtained the highest levels of user engagement with COVID-19 media and social media articles containing misinformation. This Boolean search string returned over 700 thousand social media posts which were initially filtered by user engagement and reach to provide the most widely shared and viewed posts. Two independent coders (SP and KdG) screened top posts and excluded posts that failed criteria (1-4) above. Some posts had relatively low levels of engagement, yet were included because they repeatedly appeared in different formats across different outlets and were thus deemed to be influential on social media. A set of five final posts were obtained for the US and the UK. For instance, misinformation selected to be shown to the US sample included a post falsely claiming that a COVID-19 vaccine will alter DNA in humans, while that in the UK included a post falsely claiming that COVID-19 vaccine will cause 97% of recipients to become infertile.

(*Control Set*) The aim of exposing people to factual coronavirus vaccine information is to serve as a control against the treatment exposure of misinformation since (a) exposure to *any* information can in principle cause respondents to change their vaccine inclination (control information therefore controls for other elements of our survey), and (b) respondents may misreport post-exposure vaccine intent due to recall bias or other between-conditions difference. Factual information was also obtained via Meltwater® using the same Boolean search term above, but excluding the last clause containing misinformation-specific search keys. Over 100 posts were returned that were filtered to a final set of five. Information was often from authoritative sources (or otherwise referenced to authoritative sources) such as vaccine groups and scientific organisations. We ensured that these five posts were not overtly pro-vaccination, and did not reference anti-vaccination campaigns or materials. For instance: information presented included information on: the current state of coronavirus vaccine trials; the importance of a vaccine to get out of the COVID-19 pandemic; and how a candidate vaccine generates good immune response.

See Appendix B for more details regarding image selection for both treatment and control image sets. These final image sets are shown in Figure 1 (misinformation) and Figure 2 (factual information).

### Statistical Methods

Bayesian statistical modelling is used to answer all quantitative research questions. Relevant statistics for parameters of interest (percentages, odds-ratios and log odds-ratios) are reported as a mean estimate (the effect size) with corresponding 95% percentile intervals (PI) (that is, values at 2.5% and 97.5% percentiles) to indicate credible values of the statistic. A percentage (%) or log odds ratio (log OR) is deemed “significant” if the 95% PI excludes zero, and an odds ratio (OR) is deemed “significant” if the 95% PI exclude one.

#### Measuring vaccine intent and changes in intent

Exposure to misinformation on an online social media platform is a complex system to model. There are many confounding factors regarding: psychological, socio-economic and cultural characteristics of consumers of information—traits that determine consumption of both offline and online media; characteristics of platforms serving this information including the algorithms which deliver information on social media can be opaque black-boxes; and characteristics of the information itself, such as style and semantic content, or the identity of the entity sharing the information, such as friends, family, celebrities, or organisations. These factors make it difficult to determine the “causal” impact of exposure to any piece of information (Bursztyn et al. 2020). However, we can control for some of these effects by pursuing a randomised-controlled-trial strategy to make some progress towards measuring causal impact (Hornsey 2020). (See Figure C1 in Appendix C which shows a causal diagram that simplifies the online information dynamics, and how they may influence personal beliefs, such as vaccine intent. The figure also depicts how conducting an “exposure”, where respondents are divided into a treatment group that is shown COVID-19 vaccine misinformation and a control group that is shown factual information, allows for a causal measurement of the drop in vaccine intent in response to the exposure to misinformation.)

Throughout the analysis, individuals’ responses to the vaccination intent survey questions (see *Methods: Survey Design* (1) and (2)) are modelled as ordinal variables. Vaccine intent of a respondent before (after) any intervention is modelled as *Y*_*PRE*_ ∈ {1,2,3,4} (*Y*_*POST*_ ∈ {1,2,3,4}), where 1 corresponds to the survey response “yes, definitely” and 4 corresponds to “no, definitely not” so that we have the ordering 1 < 2 < 3 < 4. Individuals are assigned an intervention group *G* ∈ {*T, C*}: treatment (*T)* or control (*C*).

To judge the impact of exposure to factual information or misinformation, we define a statistic, *Δ*, that measures the net change of respondents providing a specific response *a* ∈ {1,2,3,4}, given their specific exposure,

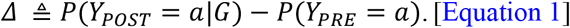

When *G* = *T* (*C*), then this statistic measures the effect of exposure to misinformation (factual information).

#### Response and explanatory variables

##### Vaccination intent

Individual variation in vaccine intent *Y*_*PRE*_ and *Y*_*POST*_ is modelled via ordered logistic regression models (McElreath 2020). That is, *Y* ∼ *OrderedLogit*(*β, α*) where *α* is an ordered vector of length 3 that encodes the intercepts of the cumulative distribution for *Y*(*Y*_*PRE*_, *Y*_*POST*_), and *β* is a placeholder for the set of predictor variables included in the regression. *Y*_*PRE*_ as a predictor variable—for susceptibility models with outcome *Y*_*POST*_—is treated as an ordinal variable containing 4 categories indexed {1,2,3,4}, where the overall slope is separated from the contribution of every additional category level by considering the parameter *δ*_*j*_ such that *δ*_*j*_ > 0 and 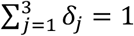, and the total slope for category *Y*_*PRE*_ (*i*) can be written as 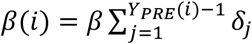. Since *δ*is a 3-simplex (the first category has a reference contribution of 0) we place a weak Dirichlet prior on it *δ* ∼ *Dirichlet*(1, 1, 1).

##### Individual level characteristics

Individual characteristic data (for the respondent *i*) is modelled as a sum of slopes across their demographic categories. For example, if a model used individuals’ age and gender then the parameters associated with the explanatory data is specified as, *β*(*i*) = *β*_*AGE*(*i*)_ + *β*_*GENDER*(*i*)_. Age (AGE), gender (GEN), highest education qualification received (EDU), (pre-pandemic) employment status (EMP), religion (REL), political affiliation (POL), ethnicity (ETH), income (INC), social media use (SOCIAL), reasons for vaccination intent (REASON), and sources of information that are trusted (TRUST) are modelled as categorical data.

All model specifications, including likelihood and prior choices are outlined below—with sensibly regularising priors chosen to prevent overestimation of effects. Table 3 illustrates the selection of explanatory variables used in each model.

**Table 3:**
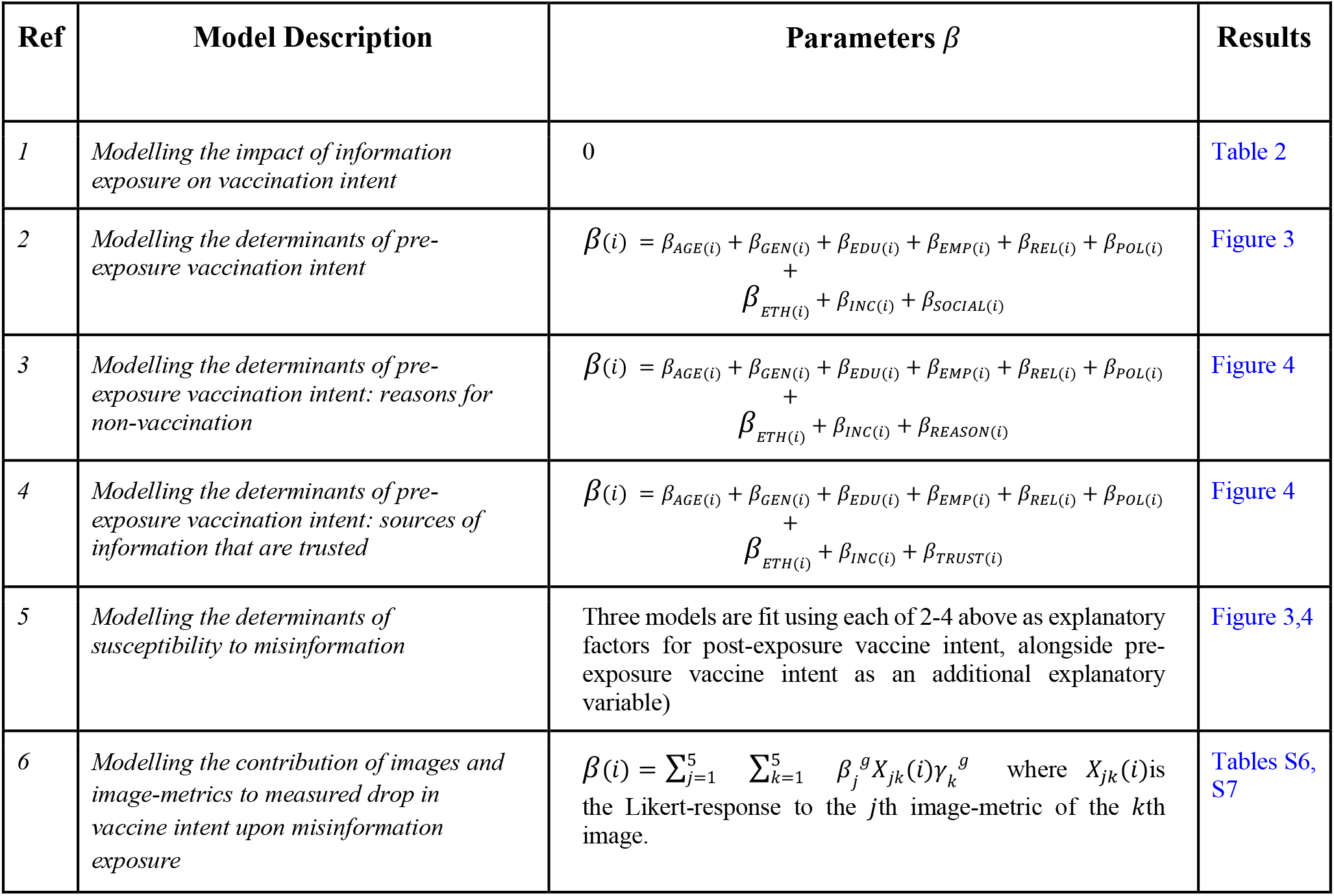
Bayesian ordinal logistic regression models for vaccine intent in the main study. The full specification of each model, and of models with outcomes other than vaccine intent, can be found in the main text.

| Ref | Model Description | Parameters $\beta$ | Results |
| --- | --- | --- | --- |
| 1 | *Modelling the impact of information exposure on vaccination intent* | 0 | Table 2 |
| 2 | *Modelling the determinants of pre-exposure vaccination intent* | $\beta(i) = \beta_{AGE(i)} + \beta_{GEN(i)} + \beta_{EDU(i)} + \beta_{EMP(i)} + \beta_{REL(i)} + \beta_{POL(i)} + \beta_{ETH(i)} + \beta_{INC(i)} + \beta_{SOCIAL(i)}$ | Figure 3 |
| 3 | *Modelling the determinants of pre-exposure vaccination intent: reasons for non-vaccination* | $\beta(i) = \beta_{AGE(i)} + \beta_{GEN(i)} + \beta_{EDU(i)} + \beta_{EMP(i)} + \beta_{REL(i)} + \beta_{POL(i)} + \beta_{ETH(i)} + \beta_{INC(i)} + \beta_{REASON(i)}$ | Figure 4 |
| 4 | *Modelling the determinants of pre-exposure vaccination intent: sources of information that are trusted* | $\beta(i) = \beta_{AGE(i)} + \beta_{GEN(i)} + \beta_{EDU(i)} + \beta_{EMP(i)} + \beta_{REL(i)} + \beta_{POL(i)} + \beta_{ETH(i)} + \beta_{INC(i)} + \beta_{TRUST(i)}$ | Figure 4 |
| 5 | *Modelling the determinants of susceptibility to misinformation* | Three models are fit using each of 2-4 above as explanatory factors for post-exposure vaccine intent, alongside pre-exposure vaccine intent as an additional explanatory variable) | Figure 3,4 |
| 6 | *Modelling the contribution of images and image-metrics to measured drop in vaccine intent upon misinformation exposure* | $\beta(i) = \sum_{j=1}^5 \sum_{k=1}^5 \beta_j^g X_{jk}(i) \gamma_k^g$ where $X_{jk}(i)$ is the Likert-response to the $j$th image-metric of the $k$th image. | Tables S6, S7 |

#### Model Specifications

##### Model for impact of information exposure on vaccine intent (Ref 1, Table 3)

Likelihood distributions: The analysis is conditioned on the groups (treatment or control) modelled independently.

1. *Y*_*PRE*_(*i*)|*G*(*i*) ∼ *OrderedLogit*(0, (*α*_1_^*PRE*^, *α*_2_^*PRE*^, *α*_3_^*PRE*^))
2. *Y*_*POST*_(*i*)|*G*(*i*) ∼ *OrderedLogit*(0, (*α*_1_^*POST*^, *α*_4_^*POST*^, *α*_3_^*POST*^))

where *y* ∼ *OrderedLogit*(*β*, (*α*_1_, *α*_1_, … *α*_*k*−1_)) implies that *log*(*P*(*y* ≤ *j*)/(1 − *P*(*y* ≤ *j*))) = *α*_*j*_ – *β*for *j* ∈ {1,2, … *k* − 1}

Distributions on priors:

1. *α*_1_^*PRE*^, *α*_2_^*PRE*^, *α*_3_^*PRE*^, *α*_1_^*POST*^, *α*_2_^*POST*^, *α*_3_^*POST*^ ∼ *Normal*(*μ, σ*)
2. *μ* ∼ *Normal*(0,1)
3. *σ* ∼ *Exponential*(1)

Constraints:

1. 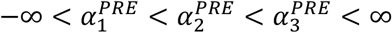
2. 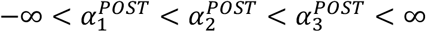

Results: Table 2

##### Models for determinants of pre-exposure vaccine intent (Ref 2-4, Table 3)

Likelihood distribution: The analysis is not conditioned on groups.

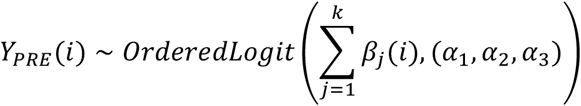

where *β*_*j*_(*i*) indicates the contribution of the *j*^th^ of *k* determinants of the *i*^th^ respondent. For instance, when doing the socio-demographic determinants analysis, the entire slope is given by:

*β*(*i*) = *β*_*AGE*(*i*)_ + *β*_*GEN*(*i*)_ + *β*_*EDU*(*i*)_ + *β*_*EMP*(*i*)_+ *β*_*REL*(*i*)_ + *β*_*POL*(*i*)_+ *β*_*ETH*(*i*)_ + *β*_*INC*(*i*)_ + *β*_*SOCIAL*(*i*)_. For analysing reasons of hesitancy, since we still control for socio-demographics, *β*_*SOCIAL*(*i*)_ is replaced by *β*_*REASON*(*i*)_, and similarly for sources of information that are trusted, *β*_*SOCIAL*(*i*)_ is replaced by *β*_*SOCIAL*(*i*)_. Distributions on priors:

1. *α*_1_, *α*_2_, *α*_3_ ∼ *Normal*(0,1)
2. *β*_*j*_ ∼ *Normal*(0,1)

Constraint: −∞ < *α*_1_ < *α*_2_ < *α*_3_ < ∞

Results: Figures 3, 4; Tables S3, S4, S5

##### Models for determinants of susceptibility to misinformation as measured by drop in vaccine intent upon exposure (Ref 5, Table 3)

Likelihood distribution: The analysis is conditioned on the groups (treatment or control) modeled independently.

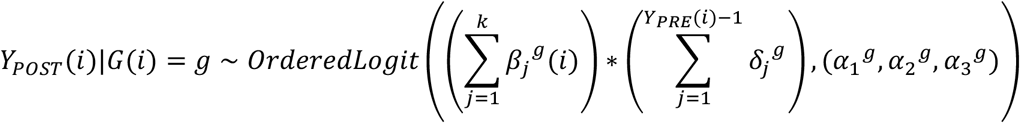

where *β*_*j*_^*g*^(*i*) indicates the contribution of the *j*^th^ of *k* determinants of the *i*^th^ respondent. Distributions on priors:

1. *α*_1_^*T*^, *α*_2_^*T*^, *α*_3_^*T*^, *α*_1_^*C*^, *α*_2_^*C*^, *α*_3_^*C*^ ∼ *Normal*(0,1)
2. *β*_*j*_^*T*^, *β*_*j*_^*C*^ ∼ *Normal*(0,1)
3. *δ*^*T*^, *δ*^*C*^ ∼ *Dirichlet*(1, 1, 1)

Constraints:

1. 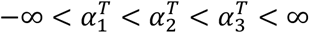
2. 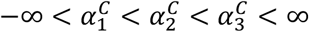

Results: Figures 3, 4; Tables S3, S4, S5

##### Model for contribution of images and image-metrics to measured drop in vaccine intent upon misinformation exposure (Ref 6, Table 3)

Likelihood distribution: The analysis is conditioned on the groups (treatment or control) modeled independently.

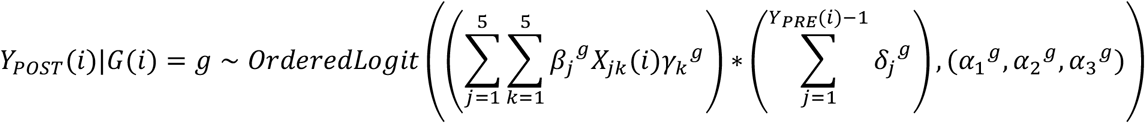

where *X*_*jk*_(*i*) indicates the Likert-response of the *i*^th^ respondent to the *j*^th^ metric of the *k*^th^ image. Here, we assume a signed response *X*_*jk*_ (*i*) ∈ {−2, −1,0,1,2}corresponding to the negative and positive ratings of a 5-level Likert scale. This allows us to gauge both (a) which images have the most impact (from γ) and (b) which image metrics/features have the most impact (from β).

Distribution on priors:

1. *α*_1_^*T*^, *α*_2_^*T*^, *α*_3_^*T*^, *α*_1_^*C*^, *α*_2_^*C*^, *α*_3_^*C*^ ∼ *Normal*(0,1)
2. *β*_*j*_ ^*T*^, *β*_*j*_^*C*^ ∼ *Normal*(0,1)
3. *γ*^*T*^, *γ*^*C*^ ∼ *Dirichlet*(1, 1, 1, 1, 1)
4. *δ*^*T*^, *δ*^*C*^ ∼ *Dirichlet*(1, 1, 1)

Constraints:

1. 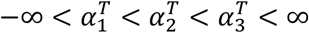
2. 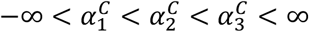

Results: Tables S6, S7

##### Model for self-reported image-metrics

For each of the 5 images shown, respondents were asked to report perceptions of the image across 5 dimensions (see *Methods: Survey Design*) on a 5-level Likert scale. This model treats the response for each image and image-metric as an ordinal variable.

Likelihood distribution: The analysis is conditioned on the groups (treatment or control), images and image-metrics modelled independently.

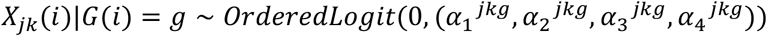

where *X*_*j k*_ (*i*) indicates the Likert-response of the *i*^th^ respondent to the *j*^th^ metric of the *k*^th^ image. Here, we assume a categorical response *X*_*j k*_ (*i*) ∈ {1,2,3,4,5} corresponding to the ordinal ratings of a 5-level Likert scale.

Distribution on priors: *α*_1_^*jkg*^, *α*_2_^*jkg*^, *α*_3_^*jkg*^, *α*_4_^*jkg*^ ∼ *Normal*(0,1)

Constraints: −∞ < *α*_1_^*jkg*^ < *α*_2_^*jkg*^ < *α*_3_^*jkg*^ < *α*_4_^*jkg*^ < ∞

Results: Figure 5

##### Model for evidence of filter-bubble effects of (mis)information exposure with regards to vaccine intent

After exposure to (mis)information, respondents were asked if they had seen “similar” information on social media recently (see *Methods: Survey Design*), to which they could respond with “yes”, “no”, or “do not know”. This model treats this response as an ordinal variable.

Likelihood distribution: The analysis is conditioned on the groups (treatment or control) modeled independently. Here, we do not treat *Y*_*PRE*_ as ordinal, to capture any U-shaped effects. Let *S* ∈ {1,2}refer to whether the respondents had “seen similar content online in the last month on social media,” where 1 implies “yes”, 2 implies “no” and “do not know”s were ignored.

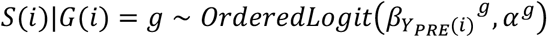

Distributions on priors:

1. *α*^*T*^, *α*^*C*^ ∼ *Normal*(0,1)
2. *β*_1_^*T*^, *β*_2_^*T*^, *β*_3_^*T*^, *β*_4_^*T*^, *β*_1_^*C*^, *β*_2_^*C*^, *β*_3_^*C*^, *β*_4_^*C*^ ∼ *Normal*(0,1)

Results: Table S8; Figure S3

#### Model Inference

Model inference was performed via Hamiltonian Monte Carlo (HMC) with the NUTS sampler using the python implementation of Stan (Carpenter et al. 2017) called pystan (Stan Development Team 2018). Samples from the posterior distribution of the model parameters were collected from 4 chains and 2000 iterations (i.e. 4000 samples excluding warm-up) after ensuring model convergence, with the potential scale reduction factor satisfying *R* ≤ 1.1 for all model parameters (Gelman & Rubin 1992). Relevant statistics for parameters of interest (coefficients, contrasts, odds-ratios and percentages) were extracted from the samples, and all results report the mean estimate—to judge magnitude of the effect—alongside 95% percentile intervals (PI) (i.e. values at 2.5% and 97.5% percentiles) to indicate credible values of the statistic—to judge significance of the effect.

## Supporting information

STROBE checklist

## Data Availability

All data for this study will be made publicly upon successful acceptance and peer review.

## Acknowledgments

This project was funded by the UN’s Verified Initiative. The funders had no role in data collection or study design. The Vaccine Confidence Project(tm) would like to acknowledge its long partnership and collaboration with ORB (Gallup) International who collected all data for this study.

## Ethical Approval

Approval for this study was obtained by the LSHTM ethics committee on 15 June 2020 with reference 22130.

## Author contributions

All authors contributed to questionnaire design with AdF obtaining ethical approval for the study via the LSHTM ethics committee. SL and AdF designed the statistical analyses. SL performed all model calculations and inference and created all publication figures. SP, KdG, and SL performed image selection. AdF and SL wrote the final manuscript with input from all authors. All authors contributed to the interpretation of the results. AdF and HL supervised the project.

## Data availability

The data are not public, but researchers can apply to use the resource.

## Code availability

The code for this project will be made available at https://github.com/sloomba/covid19-misinfo/.

