## Appendix for "Measuring the Impact of Exposure to COVID-19 Vaccine Misinformation on Vaccine Intent in the UK and US"

#### Appendix A: Socio-demographic Recoding

To reduce the number of socio-demographic categories, and make them comparable across the UK and US, we have recoded some of the respondent characteristics as described below.

| Factor | Value | Recode |
|---|---|---|
| Age | Numeric from 18 to 120 | 18-24, 25-34, 35-44, 45-54, 55-64, 65+ |
| Gender | Male, Female | |
| | 1. Other<br>2. Prefer not to answer | Other |
| Education (UK only) | No academic or professional qualifications | Level-0 |
| | 1. 0-4 GCSE, O-level or equivalents<br>2. 5+ GCSE, O-level, 1 A level, or equivalents | Level-1 |
| | 2+ A levels, or equivalents | Level-2 |
| | Undergraduate degree | Level-3 |
| | Postgraduate degree or other professional degrees | Level-4 |
| | 1. Apprenticeship<br>2. Other (e.g. vocational, foreign qualifications)<br>3. Prefer not to answer | Other |
| Education (US only) | 1. No academic or professional qualifications<br>2. Nursery or preschool through grade 12 | Level-0 |
| | High school diploma or GED | Level-1 |
| | 2-year college degree | Level-2 |
| | 4-year college degree | Level-3 |
| | Postgraduate degree or other professional degrees | Level-4 |
| | 1. Other<br>2. Prefer not to answer | Other |
| Employment | 1. Working full-time (include self-employed)<br>2. Working part-time (include self-employed) | Employed |
| | Unemployed, Student, Retired | |
| | 1. Looking after family or home<br>2. Long-term sick or disabled<br>3. Prefer not to answer | Other |

| | | |
|---|---|---|
| Religion | 1. Roman Catholic<br>2. Protestant<br>3. Other Christian | Christian |
| | Jewish, Muslim | |
| | Atheist or agnostic | Atheist |
| | 1. Hindu<br>2. Buddhist<br>3. Other<br>4. Prefer not to answer | Other |
| Political Affiliation (UK only) | Conservative, Labour, Liberal Democrat, SNP | |
| | 1. Other<br>2. Don't know<br>3. Prefer not to answer | Other |
| Political Affiliation (US only) | Republican, Democrat | |
| | 1. Independent<br>2. Don't know<br>3. Prefer not to answer | Other |
| Ethnicity (UK only) | 1. White: English/Welsh/Scottish/Northern Irish/British<br>2. White: Irish<br>3. White: Other white background | White |
| | 1. Black, African, Caribbean, or Black British<br>2. White and Black Caribbean, or White and Black African | Black |
| | 1. Asian or Asian British: Indian<br>2. Asian or Asian British: Pakistani<br>3. Asian or Asian British: Bangladeshi<br>4. Asian or Asian British: Chinese<br>5. Asian or Asian British: Other<br>6. White and Asian | Asian |
| | 1. Other<br>2. Prefer not to answer | Other |
| Ethnicity (US only) | Non-Hispanic White | White |
| | Hispanic, Asian | |
| | Black or African American | Black |
| | 1. American Indian or Alaska Native<br>2. Native Hawaiian or Pacific Islander<br>3. Other<br>4. Prefer not to answer | Other |

| | | |
|---|---|---|
| Income<br>(UK only) | Under £15,000 | Level-0 |
| | £15,000 - £24,999 | Level-1 |
| | £25,000 - £34,999 | Level-2 |
| | 1. £35,000 - £44,999<br>2. £45,000 - £54,999 | Level-3 |
| | 1. £55,000 - £64,999<br>2. £65,000 - £74,999<br>3. £75,000 - £84,999<br>4. £85,000 - £94,999<br>5. £95,000 or over | Level-4 |
| | Prefer not to answer | Other |
| Income<br>(US only) | Under $15,000 | Level-0 |
| | 1. $15,000 - $24,999<br>2. $25,000 - $34,999 | Level-1 |
| | 1. $35,000 - $44,999<br>2. $45,000 - $54,999 | Level-2 |
| | 1. $55,000 - $64,999<br>2. $65,000 - $74,999<br>3. $75,000 - $84,999<br>4. $85,000 - $94,999 | Level-3 |
| | $95,000 or over | Level-4 |
| | Prefer not to answer | Other |

#### Appendix B: Image Selection

##### Control Images

| Image | Engagement[1] | Reach[2] | Source | URL[3] |
|---|---|---|---|---|
| 1 | N.A. (Qualitative selection) | N.A. | Vaccine Safety Net / The Guardian | https://twitter.com/VaccineSafetyN/status/1288798996878819328 |
| 2 | N.A. (Retweet) | 51.5k | Bill Gates / Coalition for Epidemic Preparedness Innovations | https://twitter.com/CEPIvaccines/status/1293235060070645762 |
| 3 | 75k | 630k | University of Oxford | https://twitter.com/UniofOxford/status/1285210154984710145 |
| 4 | 1.28k | 123k | Gavi, the Vaccine Alliance | https://twitter.com/gavi/status/1291713207820836865 |
| 5 | 81.8k | 121m | Barack Obama / The Atlantic | https://twitter.com/BarackObama/status/1286729522956689410 |

##### Treatment Images

| Image | Engagement | Reach | Source | URL |
|---|---|---|---|---|
| | | UK | | |
| 1 | 1.59k | 1.5m | Twitter user | https://twitter.com/PrisonPlanet/status/1262701005382340610 |
| 2 | 27 | 19.6k | News from the Perimeter (Twitter) | https://twitter.com/PerimeterNews/status/1286439514940960769 |
| 3 | 11 | 1.49k | Twitter user | https://twitter.com/Prometheous2020/status/1297659974622023681 |
| 4 | N.A. | 32.5k | Twitter user | https://twitter.com/Jimcorrsays/status/1296780071374598144 |
| 5 | 6.95k | 336k | David Icke (Twitter) | https://twitter.com/davidicke/status/1273330307626864642 |
| | | US | | |
| 1 | 27 | 19.6k | News from the Perimeter (Twitter) | https://twitter.com/PerimeterNews/status/1286439514940960769 |
| 2 | 11 | 1.49k | Twitter user | https://twitter.com/Prometheous2020/status/1297659974622023681 |
| 3 | 25.1k | 1.41k | Twitter user | https://twitter.com/uTobian/status/1293721217791217665 |

[1] Measures the number of likes and retweets
[2] Measures the number of followers and thus potential audience size
[3] Links accessed on 25th August 2020

| | | | | |
|---|---|---|---|---|
| 4 | N.A. (Qualitative selection) | N.A. | Larry Cook (Facebook) | https://www.facebook.com/LarryCook333 |
| 5 | 28.2k | N.A. | Robert F. Kennedy Jr (Instagram) | https://www.instagram.com/p/CDsLREtHQog/ |

#### Appendix C: Statistical Modelling

##### *Impact of misinformation exposure on vaccine intent, relative to impact of factual information*

Respondents may change their intent to vaccinate when exposed to *any* kind of information. Therefore, we performed a hierarchical Bayesian ordered logistic regression (see model specification below) to model the distribution of post-exposure intent $Y_{POST}$, given pre-exposure intent $Y_{PRE}$ and the group $G \in \{T, C\}$which the respondent was assigned to. This is precisely the functional relationship expressed by the causal diagram in Figure C1. Following which, we can estimate the posterior distribution of the causal impact as measured by the risk difference:

$$\Delta_{RD} \triangleq P(Y_{POST} = a|Y_{PRE} = b, G = T) - P(Y_{POST} = a|Y_{PRE} = b, G = C), \qquad \text{[Equation 2]}$$

where $a, b \in \{1,2,3,4\}$refers to a pair of the four vaccine intents. This statistic responds to the following query: **of those with pre-exposure intent *b*, what proportion of people change their intent to *a* when exposed to misinformation, relative to when exposed to factual information?** This is the statistic depicted in Figure S2 of Appendix D, and quoted as $\Delta_{RD}$ in Tables S1, S2 of Appendix D.

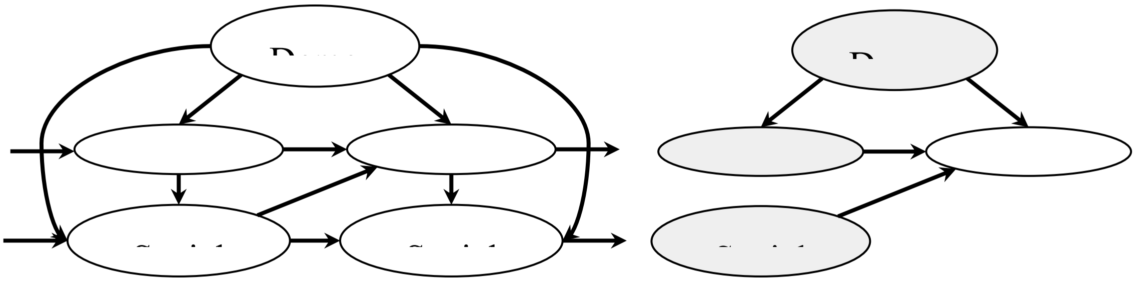

Figure C1 **(a) Causal diagram indicating the influence of socio-demographic and other determinants of vaccine intent, which is dynamically influenced by exposure to social media content in the real world.** *t* refers to a particular time-point. Evidently, demographics (among other characteristics) will influence an individual's vaccine intent at all time-points. Moreover, it will also influence the kind of social media content they are exposed to, as per the algorithms of the social media platform they use. These algorithms may also be taking the individual's vaccine intent as input (likely latent in the content they "like" or "share" on the platform) and expose them to more content conditioned on that. Other possible confounders, such as different sources of information, have been ignored here for simplicity. **(b) Causal diagram of the experiment conducted in this study, which attempts to model exposure to information in the form of a randomised controlled trial.** This removes the confounding influence of demographics and vaccine intent on the social media content that individuals might see in the real world.

##### *Model Specification*

Outcome distribution: The analysis is conditioned on the groups (treatment or control) with hierarchical priors.

$$Y_{POST}(i)|G(i) = g \sim OrderedLogit\left(\beta^g * \left(\sum_{j=1}^{Y_{PRE}(i)-1} \delta_j{}^g\right), (\alpha_1{}^g, \alpha_2{}^g, \alpha_3{}^g)\right)$$

Distributions on priors:

1. $\alpha_1{}^T, \alpha_2{}^T, \alpha_3{}^T, \alpha_1{}^C, \alpha_2{}^C, \alpha_3{}^C \sim Normal(\mu_\alpha, \sigma_\alpha)$
2. $\beta^T, \beta^C \sim Normal(\mu_\beta, \sigma_\beta)$
3. $\delta^T, \delta^C \sim Dirichlet(\sigma_\delta * \rho_\delta)$
4. $\mu_\alpha, \mu_\beta \sim Normal(0,1)$
5. $\sigma_\alpha, \sigma_\beta, \sigma_\delta \sim Exponential(1)$

6. $\rho_\delta \sim Dirichlet(1, 1, 1)$

Constraints:

1. $-\infty < {\alpha_1}^T < {\alpha_2}^T < {\alpha_3}^T < \infty$
2. $-\infty < {\alpha_1}^C < {\alpha_2}^C < {\alpha_3}^C < \infty$

Results: Tables S1, S2; Figures S1, S2

### Appendix D: Supplementary Tables and Figures

**UNITED KINGDOM**

| Group | POST / PRE | Yes, definitely | Unsure, lean yes | Unsure, lean no | No, definitely not |
|---|---|---|---|---|---|
| Treatment | Yes, definitely | 80.8 (78.8, 82.6) | 18.0 (16.2, 19.8) | 1.1 (0.9, 1.4) | 0.2 (0.1, 0.2) |
| | Unsure, lean yes | 15.1 (13.2, 17.3) | 61.8 (59.1, 64.6) | 19.5 (17.4, 21.7) | 3.6 (2.8, 4.5) |
| | Unsure, lean no | 1.4 (1.0, 1.8) | 19.3 (15.5, 23.4) | 47.2 (43.0, 51.3) | 32.2 (27.2, 37.5) |
| | No, definitely not | 0.1 (0.1, 0.1) | 1.6 (0.9, 2.5) | 10.6 (6.8, 15.0) | 87.7 (82.5, 92.1) |
| Control | Yes, definitely | 91.3 (88.8, 93.5) | 8.5 (6.4, 10.9) | 0.2 (0.1, 0.3) | 0.0 (0.0, 0.0) |
| | Unsure, lean yes | 17.1 (13.4, 21.2) | 74.4 (69.6, 78.8) | 7.7 (5.3, 10.5) | 0.8 (0.4, 1.3) |
| | Unsure, lean no | 0.9 (0.5, 1.5) | 31.0 (22.5, 40.2) | 53.1 (44.3, 62.0) | 14.9 (8.9, 22.1) |
| | No, definitely not | 0.1 (0.0, 0.1) | 3.3 (1.3, 6.5) | 25.9 (15.5, 37.4) | 70.7 (57.4, 82.9) |
| $\Delta_{RD}$ | Yes, definitely | **-10.5** (-13.4, -7.4) | **9.4** (6.4, 12.2) | **0.9** (0.7, 1.2) | **0.1** (0.1, 0.2) |
| | Unsure, lean yes | -2.0 (-6.4, 2.3) | **-12.6** (-17.7, -7.2) | **11.8** (8.3, 15.2) | **2.8** (1.8, 3.9) |
| | Unsure, lean no | 0.5 (-0.3, 1.1) | **-11.7** (-21.8, -2.3) | -6.0 (-15.9, 3.9) | **17.2** (8.8, 25.3) |
| | No, definitely not | 0.0 (-0.1, 0.1) | -1.7 (-5.0, 0.5) | **-15.3** (-27.2, -3.8) | **17.0** (3.6, 30.9) |

**UNITED STATES**

| Group | POST / PRE | Yes, definitely | Unsure, lean yes | Unsure, lean no | No, definitely not |
|---|---|---|---|---|---|
| Treatment | Yes, definitely | 79.5 (77.2, 81.7) | 18.7 (16.7, 20.8) | 1.6 (1.3, 1.9) | 0.3 (0.2, 0.4) |
| | Unsure, lean yes | 19.2 (16.9, 21.5) | 57.3 (54.4, 59.9) | 19.1 (17.0, 21.4) | 4.4 (3.5, 5.4) |
| | Unsure, lean no | 2.7 (2.1, 3.4) | 24.8 (21.3, 28.3) | 44.1 (40.5, 47.8) | 28.4 (24.5, 32.7) |
| | No, definitely not | 0.2 (0.2, 0.3) | 2.8 (2.1, 3.8) | 14.3 (11.5, 17.2) | 82.7 (79.0, 86.1) |
| Control | Yes, definitely | 88.1 (84.9, 91.1) | 10.8 (8.2, 13.8) | 0.9 (0.5, 1.3) | 0.2 (0.1, 0.2) |
| | Unsure, lean yes | 27.6 (22.9, 32.8) | 55.8 (50.6, 60.9) | 13.7 (10.5, 17.4) | 2.9 (1.8, 4.2) |
| | Unsure, lean no | 3.7 (2.3, 5.5) | 29.5 (23.1, 36.4) | 43.9 (37.3, 50.6) | 22.9 (16.9, 29.7) |
| | No, definitely not | 0.3 (0.1, 0.5) | 3.3 (1.9, 5.2) | 16.6 (11.6, 22.0) | 79.8 (73.2, 85.9) |
| $\Delta_{RD}$ | Yes, definitely | **-8.7** (-12.3, -4.7) | **7.8** (4.3, 11.2) | **0.7** (0.2, 1.2) | **0.1** (0.0, 0.2) |
| | Unsure, lean yes | **-8.5** (-14.2, -3.0) | 1.5 (-4.3, 7.4) | **5.4** (1.2, 9.3) | 1.6 (-0.0, 3.0) |
| | Unsure, lean no | -1.0 (-3.0, 0.6) | -4.8 (-12.3, 2.6) | 0.2 (-7.1, 7.8) | 5.6 (-2.3, 13.1) |
| | No, definitely not | -0.1 (-0.3, 0.1) | -0.5 (-2.6, 1.2) | -2.3 (-8.5, 3.6) | 2.9 (-4.4, 10.6) |

Table S1 **Exposure to COVID-19 vaccine misinformation reduces inclination to vaccinate to "protect themselves", relative to factually correct information.** Rows refer to pre-exposure vaccine intent and columns refer to post-exposure vaccine intent. Values indicate mean estimate of percentages, with 95% percentile intervals indicated in parentheses. Underlined values refer to the probability of sticking with the pre-exposure vaccine intent, for either the treatment or control groups. $\Delta_{RD}$ refers to a risk-difference measure of causal impact of exposure to misinformation relative to exposure to factually correct information, as described in Equation 2 in Appendix C. For example in the UK, of those who said they would "definitely" get vaccinated, about 10.5% picked more hesitant options upon exposure to misinformation relative to exposure to factually correct information. This number was 8.7% in the US. Evidently, the impact of misinformation appears to be higher in the UK, with a significant 17% more people sticking with "No, definitely not" after exposure to misinformation.

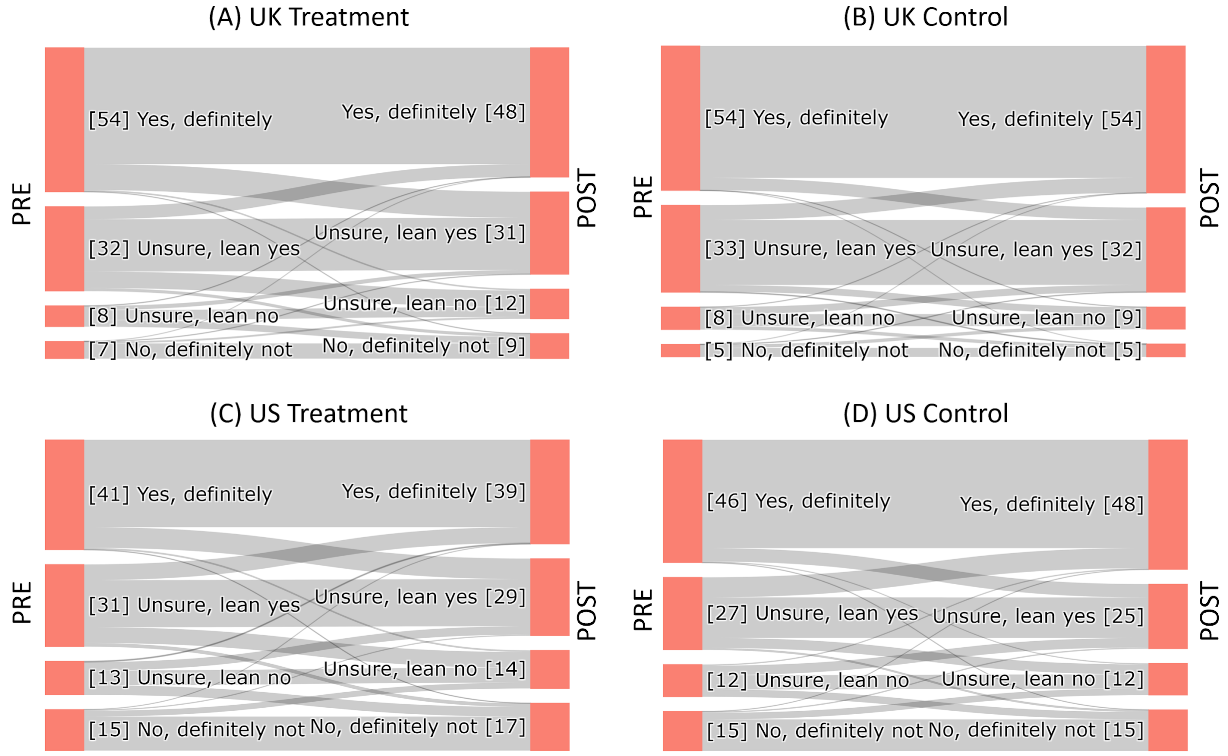


Figure S1 **Sankey diagrams illustrate how individuals change their intent to vaccinate to protect themselves before (PRE) and after (POST) exposure to factually incorrect (treatment) or correct (control).** The percentage of respondents in the UK (A, B) and US (C, D) replying to each vaccination intent category pre- (red bars on left) and post-exposure(red bars on right) is shown with individual shifts in sentiment, represented by gray flows. Although individuals can obtain a higher or lower sentiment when exposed to the treatment (A, C) or control images (B, D), there is an overall shift towards COVID-19 vaccine hesitancy for treatment groups but no overall shift in hesitancy for control groups.

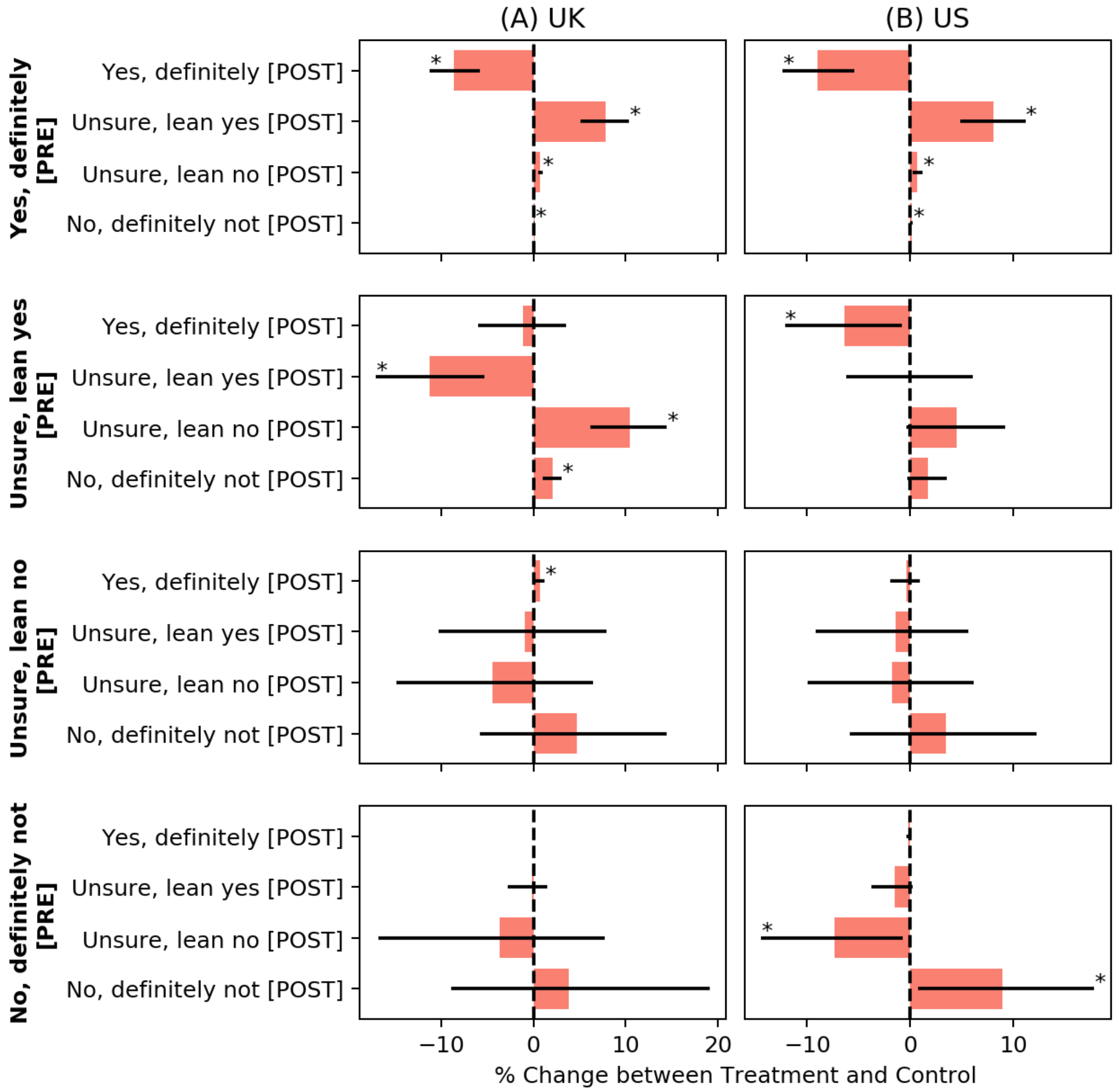


Figure S2 **Exposure to misinformation induces vaccine hesitancy among all positive pre-exposure response groups.** The differences in the percentage of respondents falling into a response category for the treatment group versus the control group after exposure (red bars; see Equation 2, Appendix C) for the UK (A) and USA (B), when asked about getting vaccinated to protect others who are at-risk. Black lines denote 95% percentile intervals and are starred (*) if this interval excludes zero. For example, in the UK, 8.6% fewer respondents in the treatment group than the control group still reported that they would “definitely” take a COVID-19 vaccine after exposure, while this number was 9.0% in the US.

| Group | PRE \ POST | Yes, definitely | Unsure, lean yes | Unsure, lean no | No, definitely not |
|---|---|---|---|---|---|
| **UNITED KINGDOM** | | | | | |
| Treatment | Yes, definitely | <u>82.1</u> (80.3, 83.9) | 16.9 (15.2, 18.5) | 0.9 (0.7, 1.1) | 0.1 (0.1, 0.1) |
| | Unsure, lean yes | 13.9 (11.8, 16.2) | <u>63.5</u> (60.6, 66.4) | 19.6 (17.3, 22.2) | 3.0 (2.2, 3.9) |
| | Unsure, lean no | 1.2 (0.8, 1.6) | 19.2 (15.2, 23.5) | <u>50.4</u> (45.8, 55.0) | 29.2 (24.2, 34.7) |
| | No, definitely not | 0.1 (0.0, 0.1) | 1.6 (0.9, 2.6) | 12.2 (7.5, 17.5) | <u>86.2</u> (79.9, 91.5) |
| Control | Yes, definitely | <u>90.7</u> (88.4, 92.8) | 9.1 (7.0, 11.4) | 0.2 (0.1, 0.3) | 0.0 (0.0, 0.0) |
| | Unsure, lean yes | 15.0 (10.9, 19.3) | <u>74.8</u> (69.6, 79.8) | 9.2 (6.3, 12.9) | 1.0 (0.5, 1.7) |
| | Unsure, lean no | 0.5 (0.2, 1.0) | 20.1 (12.5, 28.7) | <u>54.8</u> (45.4, 64.2) | 24.5 (16.4, 33.7) |
| | No, definitely not | 0.0 (0.0, 0.1) | 1.8 (0.5, 4.3) | 15.8 (6.1, 28.7) | <u>82.4</u> (67.8, 93.3) |
| $\Delta_{RD}$ | Yes, definitely | **-8.6** (-11.3, -5.8) | **7.8** (5.1, 10.4) | **0.7** (0.5, 1.0) | **0.1** (0.1, 0.1) |
| | Unsure, lean yes | -1.1 (-6.0, 3.5) | **-11.3** (-17.1, -5.3) | **10.4** (6.2, 14.4) | **2.0** (1.0, 3.1) |
| | Unsure, lean no | **0.7** (0.1, 1.2) | -0.9 (-10.3, 7.9) | -4.4 (-14.8, 6.4) | 4.7 (-5.8, 14.4) |
| | No, definitely not | 0.0 (-0.0, 0.1) | -0.2 (-2.8, 1.5) | -3.6 (-16.8, 7.7) | 3.8 (-8.9, 19.1) |
| **UNITED STATES** | | | | | |
| Treatment | Yes, definitely | 79.0 (76.8, 81.0) | 19.1 (17.3, 21.1) | 1.6 (1.3, 2.0) | 0.3 (0.2, 0.4) |
| | Unsure, lean yes | 16.9 (14.6, 19.3) | 56.7 (53.8, 59.7) | 21.1 (18.7, 23.6) | 5.2 (4.1, 6.4) |
| | Unsure, lean no | 2.3 (1.7, 2.9) | 21.9 (18.4, 25.4) | 43.3 (39.7, 47.0) | 32.6 (28.2, 37.2) |
| | No, definitely not | 0.2 (0.1, 0.2) | 2.0 (1.4, 2.8) | 10.5 (7.8, 13.5) | 87.4 (83.7, 90.7) |
| Control | Yes, definitely | 88.0 (85.0, 90.6) | 11.0 (8.6, 13.7) | 0.9 (0.6, 1.3) | 0.1 (0.1, 0.2) |
| | Unsure, lean yes | 23.2 (18.4, 28.5) | 56.7 (51.1, 62.2) | 16.6 (12.7, 20.8) | 3.5 (2.1, 5.2) |
| | Unsure, lean no | 2.6 (1.5, 4.1) | 23.3 (17.1, 30.3) | 45.0 (38.2, 52.1) | 29.1 (21.5, 37.3) |
| | No, definitely not | 0.3 (0.1, 0.5) | 3.5 (2.0, 5.6) | 17.8 (12.1, 24.2) | 78.4 (70.4, 85.4) |
| $\Delta_{RD}$ | Yes, definitely | **-9.0** (-12.4, -5.4) | **8.1** (4.9, 11.2) | **0.7** (0.2, 1.2) | **0.1** (0.0, 0.3) |
| | Unsure, lean yes | **-6.3** (-12.2, -0.8) | 0.0 (-6.2, 6.1) | 4.6 (-0.4, 9.3) | 1.7 (-0.3, 3.6) |
| | Unsure, lean no | -0.3 (-1.9, 0.9) | -1.4 (-9.2, 5.7) | -1.7 (-10.0, 6.2) | 3.5 (-5.9, 12.3) |
| | No, definitely not | -0.1 (-0.4, 0.0) | -1.5 (-3.8, 0.3) | **-7.3** (-14.5, -0.7) | **8.9** (0.8, 17.9) |

Table S2 **Exposure to COVID-19 vaccine misinformation reduces inclination to vaccinate to "protect others", relative to factually correct information.** Rows refer to pre-exposure vaccine intent and columns refer to post-exposure vaccine intent. Values indicate mean estimate of percentages, with 95% percentile intervals indicated in parentheses. <u>Underlined</u> values refer to the probability of sticking with the pre-exposure vaccine intent, for either the treatment or control groups. $\Delta_{RD}$ refers to a risk-difference measure of causal impact of exposure to misinformation relative to exposure to factually correct information, as described in Equation 2 in Appendix C. For example in the UK, of those who said they would “definitely” get vaccinated, about 8.6% picked more hesitant options upon exposure to misinformation relative to exposure to factually correct information. This number was 9.0% in the US. Contrary to the impact on intent to vaccinate to protect oneself, the impact of misinformation appears to be higher here in the US, with a significant 8.9% more people sticking with “No, definitely not” after exposure to misinformation.

| Characteristic | | UNITED KINGDOM Pre-Exposure | UNITED KINGDOM Susceptibility | UNITED STATES Pre-Exposure | UNITED STATES Susceptibility |
|---|---|---|---|---|---|
| Age | 25-34 | 1.08 (0.85, 1.36) | 0.52 (0.21, 1.08) | **1.35** (1.08, 1.67) | 1.69 (0.82, 3.08) |
| | 35-44 | 1.01 (0.78, 1.28) | 0.81 (0.33, 1.64) | 1.21 (0.94, 1.55) | **2.35** (1.08, 4.54) |
| | 45-54 | 0.97 (0.75, 1.25) | 0.88 (0.35, 1.86) | **1.60** (1.25, 2.02) | **2.62** (1.28, 4.84) |
| | 55-64 | **0.75** (0.57, 0.99) | 0.93 (0.34, 2.01) | **1.82** (1.40, 2.33) | **3.68** (1.70, 7.03) |
| | 65+ | **0.52** (0.35, 0.74) | 0.77 (0.20, 2.05) | 1.20 (0.89, 1.58) | **2.90** (1.23, 5.85) |
| Education | Level-0 | **1.85** (1.30, 2.55) | 0.93 (0.28, 2.27) | **1.40** (1.00, 1.92) | 0.72 (0.29, 1.51) |
| | Level-1 | **1.58** (1.27, 1.95) | 0.76 (0.34, 1.46) | **1.77** (1.43, 2.16) | 1.81 (0.96, 3.04) |
| | Level-2 | **1.50** (1.20, 1.88) | 1.19 (0.46, 2.56) | **1.73** (1.39, 2.14) | 1.65 (0.83, 2.92) |
| | Level-3 | 1.22 (0.99, 1.49) | 0.75 (0.33, 1.43) | **1.32** (1.08, 1.61) | 1.13 (0.60, 1.91) |
| | Other | **1.51** (1.13, 2.00) | 1.24 (0.42, 2.80) | **1.98** (1.45, 2.63) | 1.62 (0.68, 3.31) |
| Employment | Other | 0.91 (0.72, 1.13) | 0.82 (0.40, 1.53) | **1.45** (1.18, 1.78) | 1.63 (0.94, 2.61) |
| | Retired | **0.71** (0.51, 0.95) | 0.86 (0.29, 2.01) | 1.13 (0.90, 1.39) | 1.44 (0.78, 2.44) |
| | Student | 0.84 (0.59, 1.14) | 0.70 (0.21, 1.73) | **1.62** (1.18, 2.18) | 1.30 (0.50, 2.80) |
| | Unemployed | 0.99 (0.74, 1.30) | 0.76 (0.29, 1.64) | 1.10 (0.88, 1.35) | 0.93 (0.48, 1.63) |
| Ethnicity | Asian | **1.33** (1.02, 1.70) | 0.97 (0.37, 2.15) | 0.85 (0.62, 1.12) | **0.31** (0.11, 0.68) |
| | Black | **2.07** (1.44, 2.89) | 0.73 (0.26, 1.62) | **1.69** (1.39, 2.02) | 0.95 (0.54, 1.58) |
| | Hispanic | - | - | 0.86 (0.67, 1.09) | 0.76 (0.36, 1.43) |
| | Other | **2.69** (1.69, 4.06) | 1.06 (0.30, 2.73) | **2.08** (1.62, 2.64) | 0.77 (0.41, 1.37) |
| Gender | Female | **1.44** (1.25, 1.63) | 1.29 (0.75, 2.01) | **2.02** (1.78, 2.29) | 1.19 (0.82, 1.70) |
| | Other | **2.31** (1.02, 4.47) | 1.25 (0.19, 4.24) | 1.58 (0.84, 2.73) | 0.43 (0.11, 1.23) |
| Income | Level-0 | **1.56** (1.22, 1.98) | 1.20 (0.49, 2.44) | **1.72** (1.34, 2.19) | 1.04 (0.49, 1.95) |
| | Level-1 | **1.38** (1.10, 1.69) | 0.96 (0.41, 1.89) | **2.10** (1.69, 2.60) | 1.21 (0.64, 2.12) |
| | Level-2 | 1.21 (0.97, 1.51) | 0.67 (0.29, 1.33) | **1.91** (1.53, 2.36) | 0.95 (0.48, 1.71) |
| | Level-3 | 1.08 (0.88, 1.33) | 0.62 (0.27, 1.24) | **1.57** (1.28, 1.91) | 1.16 (0.59, 2.07) |
| | Other | **1.85** (1.36, 2.47) | 0.80 (0.30, 1.77) | **2.46** (1.79, 3.31) | 1.33 (0.53, 2.81) |
| Political | Democrat | - | - | **0.66** (0.56, 0.76) | 0.90 (0.55, 1.39) |
| | Labour | 0.97 (0.82, 1.13) | 1.09 (0.60, 1.84) | - | - |
| | Lib-Dem | 1.04 (0.81, 1.32) | 0.74 (0.27, 1.62) | - | - |
| | Other | **1.86** (1.54, 2.22) | **2.08** (1.13, 3.57) | **1.23** (1.05, 1.42) | 1.38 (0.89, 2.06) |

| | | | | | |
|---|---|---|---|---|---|
| | SNP | 0.88 (0.61, 1.23) | 0.75 (0.20, 2.07) | - | - |
| | Atheist | 0.89 (0.76, 1.03) | 1.09 (0.63, 1.74) | 1.09 (0.88, 1.33) | 0.77 (0.42, 1.32) |
| | Jewish | 0.63 (0.30, 1.11) | 0.87 (0.11, 3.07) | 0.90 (0.66, 1.23) | 0.50 (0.20, 1.07) |
| | Muslim | 1.30 (0.90, 1.79) | 0.89 (0.28, 2.19) | **0.55** (0.38, 0.77) | **0.34** (0.10, 0.89) |
| Religion | Other | 1.05 (0.87, 1.25) | 0.97 (0.52, 1.69) | **1.51** (1.28, 1.76) | 1.03 (0.66, 1.53) |
| | < 10 minutes | 0.92 (0.71, 1.18) | 0.77 (0.31, 1.59) | **0.64** (0.51, 0.80) | **0.57** (0.29, 0.99) |
| | 10–30 minutes | 0.91 (0.72, 1.14) | 0.72 (0.32, 1.43) | **0.82** (0.66, 1.00) | **0.60** (0.34, 0.99) |
| | 31–60 minutes | 0.94 (0.73, 1.19) | 0.71 (0.29, 1.49) | **0.70** (0.56, 0.87) | 0.82 (0.43, 1.45) |
| | 1–2 hours | 0.93 (0.72, 1.18) | 1.16 (0.49, 2.29) | 0.82 (0.64, 1.03) | 0.94 (0.47, 1.72) |
| | 2–3 hours | 0.95 (0.71, 1.23) | 1.70 (0.64, 3.74) | 0.88 (0.68, 1.13) | 1.18 (0.56, 2.18) |
| Social media usage (per day) | > 3 hours | 0.89 (0.67, 1.15) | 1.12 (0.45, 2.34) | **0.79** (0.62, 0.98) | 1.00 (0.54, 1.74) |

Table S3 **Multiple regression reveals the key determinants of intent to vaccinate—to protect themselves—before exposure and the groups most likely to be negatively impacted by misinformation.** Contribution of socio-demographic and social-media-use characteristics to pre-exposure vaccine intent and susceptibility to misinformation exposure for the UK and US. Values depict odds-ratios (OR) of being more hesitant (for pre-exposure) or more susceptible (for susceptibility)—as measured relative to the reference category of Male, 18-24, highest education, employed, Christian, White, Conservative, highest income, and no social media usage. **OR>1 indicates the group is more likely to not accept a COVID-19 vaccine than the reference group, and more likely to be susceptible to COVID-19 vaccine misinformation than the reference group.** Values in parentheses indicate 95% percentile intervals and values in **bold** indicate "statistical significance" based on them.

| | UNITED KINGDOM | | UNITED STATES | |
|---|---|---|---|---|
| **Reasons for Hesitancy** | **Pre-Exposure** | **Susceptibility** | **Pre-Exposure** | **Susceptibility** |
| Already acquired immunity | 0.26 (-0.15, 0.65) | 1.03 (-0.33, 2.47) | **0.38** (0.07, 0.69) | -0.30 (-1.07, 0.49) |
| Approval may be rushed | 0.18 (-0.06, 0.41) | **0.93** (0.07, 1.83) | 0.18 (-0.00, 0.38) | 0.35 (-0.19, 0.88) |
| Do not know | **1.22** (0.75, 1.68) | -0.08 (-1.37, 1.20) | **1.68** (1.35, 2.02) | 0.64 (-0.10, 1.40) |
| Not at risk of catching it | **0.73** (0.40, 1.05) | 0.33 (-0.77, 1.47) | **0.55** (0.27, 0.84) | 0.30 (-0.45, 1.08) |
| Other reasons | **2.13** (1.64, 2.62) | 0.86 (-0.43, 2.20) | **1.44** (1.07, 1.82) | **1.15** (0.22, 2.20) |
| Other effective treatments | 0.05 (-0.34, 0.43) | -0.45 (-1.60, 0.74) | 0.27 (-0.02, 0.55) | -0.24 (-0.94, 0.47) |
| Unsure if effective | -0.09 (-0.32, 0.15) | 0.30 (-0.53, 1.19) | 0.01 (-0.18, 0.20) | 0.05 (-0.51, 0.62) |
| Unsure if safe | **0.33** (0.09, 0.58) | 0.39 (-0.44, 1.25) | 0.14 (-0.05, 0.33) | **1.22** (0.65, 1.82) |
| Wait until others | **-0.41** (-0.67, -0.17) | -0.03 (-0.88, 0.86) | **-0.50** (-0.69, -0.31) | **-0.88** (-1.47, -0.29) |
| Won't be ill | **0.76** (0.45, 1.07) | -0.56 (-1.49, 0.38) | **0.49** (0.21, 0.78) | -0.13 (-0.85, 0.60) |

Table S4 **Concerns of vaccine safety and rushed approval are indicative of susceptibility to misinformation.** Contribution of reasons that respondents provide for not being “definitely” sure of taking a COVID-19 vaccine (to protect themselves) to the Pre-Exposure vaccine hesitancy and Susceptibility to vaccine misinformation as measured by drop in vaccine intent, for both the UK and US—after controlling for socio-demographics. Values depict odds-ratios (OR) of being more hesitant (for pre-exposure) or more susceptible (for susceptibility)—as measured relative to when the reason was not indicated for hesitancy. **OR>1 indicates that if that reason was indicated for hesitancy, then the respondent is more likely to not accept a COVID-19 vaccine.** Values in parentheses indicate 95% percentile intervals and values in **bold** indicate “statistical significance” based on them. Since reasons for hesitancy were only asked to those who did not choose “Yes, definitely” when asked if they would vaccinate to protect themselves, that analysis is conditioned to those who did not indicate “Yes, definitely” for the SELF question.

| Source of Trusted Information | UNITED KINGDOM | | UNITED STATES | |
|---|---|---|---|---|
| | Pre-Exposure | Susceptibility | Pre-Exposure | Susceptibility |
| Celebrities | **-0.65** (-1.09, -0.23) | **-1.94** (-3.48, -0.43) | **-0.43** (-0.78, -0.09) | -1.08 (-2.18, 0.02) |
| Family and friends | **0.33** (0.12, 0.53) | 0.35 (-0.39, 1.08) | 0.05 (-0.13, 0.22) | 0.13 (-0.39, 0.63) |
| Govt. Briefings | **-0.18** (-0.33, -0.03) | -0.49 (-1.07, 0.11) | - | - |
| Govt. Websites | -0.04 (-0.19, 0.11) | 0.11 (-0.45, 0.70) | **-0.28** (-0.46, -0.10) | -0.25 (-0.83, 0.32) |
| Healthcare Workers | -0.08 (-0.22, 0.06) | -0.12 (-0.67, 0.44) | 0.08 (-0.07, 0.22) | 0.05 (-0.36, 0.46) |
| International Health Authorities | **-0.25** (-0.40, -0.10) | 0.03 (-0.57, 0.60) | **-0.20** (-0.36, -0.05) | -0.38 (-0.86, 0.10) |
| National Health Authorities | **-0.22** (-0.38, -0.07) | 0.26 (-0.33, 0.85) | **-0.21** (-0.36, -0.05) | 0.08 (-0.36, 0.55) |
| Newspapers | 0.08 (-0.12, 0.28) | -0.51 (-1.25, 0.25) | **-0.22** (-0.42, -0.02) | 0.27 (-0.39, 0.95) |
| None of these | **1.38** (1.12, 1.64) | **0.74** (0.01, 1.46) | **1.44** (1.20, 1.69) | **0.71** (0.17, 1.28) |
| Other | 0.41 (-0.39, 1.18) | 0.84 (-0.91, 2.64) | **0.95** (0.43, 1.49) | 0.56 (-0.51, 1.72) |
| Radio | -0.10 (-0.35, 0.14) | 0.19 (-0.74, 1.15) | -0.16 (-0.39, 0.07) | -0.48 (-1.18, 0.25) |
| Scientists | -0.02 (-0.17, 0.13) | 0.53 (-0.02, 1.09) | 0.00 (-0.14, 0.15) | **0.45** (0.03, 0.88) |
| Search Engines | -0.07 (-0.30, 0.15) | 0.74 (-0.12, 1.63) | 0.12 (-0.07, 0.30) | 0.03 (-0.54, 0.60) |
| Social Media | 0.01 (-0.27, 0.28) | 0.13 (-0.76, 1.03) | **-0.28** (-0.50, -0.06) | **0.79** (0.08, 1.50) |
| State Govt. Briefings | - | - | -0.08 (-0.24, 0.09) | -0.04 (-0.55, 0.48) |
| Television | **-0.29** (-0.44, -0.14) | -0.17 (-0.77, 0.42) | **-0.16** (-0.32, -0.01) | -0.16 (-0.63, 0.31) |
| White House Briefings | - | - | **-0.23** (-0.41, -0.04) | 0.31 (-0.25, 0.88) |
| Work Guidelines | -0.13 (-0.39, 0.13) | -0.44 (-1.37, 0.47) | -0.20 (-0.43, 0.03) | -0.54 (-1.35, 0.26) |

Table S5 **Mistrust in mainstream sources of information is indicative of lower COVID-19 vaccine intent and higher susceptibility to misinformation.** Contribution of sources of information that people trust to the Pre-Exposure vaccine hesitancy and Susceptibility to vaccine misinformation as measured by drop in vaccine intent (to protect themselves), for both the UK and US—after controlling for socio-demographics. Values depict odds-ratios (OR) of being more hesitant (for pre-exposure) or more susceptible (for susceptibility)—as measured relative to when the source was not indicated as being trusted. **OR>1 indicates that if that source was indicated as trustworthy then the respondent is more likely to not accept a COVID-19 vaccine.** Values in parentheses indicate 95% percentile intervals and values in **bold** indicate "statistical significance" based on them.

| | Treatment | | Control | |
|---|---|---|---|---|
| **Image Characteristic** | **UK** | **US** | **UK** | **US** |
| Makes less inclined to vaccinate | **43.93** (27.04, 70.57) | **13.62** (10.07, 18.64) | 2.15 (0.99, 4.32) | **4.63** (2.52, 8.11) |
| Agree with | **2.72** (1.40, 4.82) | **1.82** (1.14, 2.78) | 1.64 (0.57, 3.86) | 2.73 (0.98, 5.69) |
| Found trustworthy | **0.35** (0.18, 0.63) | **0.45** (0.28, 0.71) | **3.05** (1.07, 7.14) | **0.50** (0.20, 0.99) |
| Likely to fact-check | **1.73** (1.32, 2.26) | **1.58** (1.22, 2.04) | **2.25** (1.39, 3.66) | **1.64** (1.11, 2.34) |
| Likely to share | **0.16** (0.10, 0.23) | **0.23** (0.17, 0.30) | **0.04** (0.02, 0.07) | **0.19** (0.12, 0.30) |

Table S6 **Image characteristics that determine their impact on the intent to vaccinate to protect themselves.** Odds-Ratios for the contribution of self-reported image characteristics to the drop in intent to vaccinate (to protect themselves) post-exposure. **OR>1 indicates that the more a respondent agreed with that image characteristic when self-reporting, the more it caused a drop in their measured vaccine intent.** Values in parentheses indicate 95% percentile intervals and values in **bold** indicate "statistical significance" based on them. For example, clearly for misinformation images, the more the respondent agreed that it (a) made them less inclined to vaccinate, and they (b) agreed with the information shown, and the (c) more likely they were to fact-check it, the more the image caused a drop in measured vaccine intent. On the other hand, the lesser the respondent found the image to be (a) trustworthy or (b) shareable, the more it contributed to drop in measured vaccine intent.

| | Treatment | | | | Control | | | |
|---|---|---|---|---|---|---|---|---|
| | UK | | US | | UK | | US | |
| | **Weight (%)** | **Rk** | **Weight (%)** | **Rk** | **Weight (%)** | **Rk** | **Weight (%)** | **Rk** |
| 1 | **36.16** (27.75, 44.86) | **1** | **34.07** (26.07, 42.27) | **1** | 20.35 (3.30, 39.87) | 3 | **27.02** (10.72, 43.37) | **1** |
| 2 | 26.19 (17.27, 35.16) | 2 | 22.30 (13.71, 30.80) | 3 | 26.13 (5.78, 46.36) | 2 | 26.46 (12.06, 41.54) | 3 |
| 3 | 9.01 (1.00, 18.10) | 4 | **5.62** (0.26, 13.74) | **5** | **9.81** (0.34, 29.07) | **5** | **8.14** (0.41, 20.49) | **5** |
| 4 | **8.13** (0.86, 17.01) | **5** | 8.45 (0.99, 16.77) | 4 | 16.73 (1.99, 35.39) | 4 | 11.52 (0.54, 31.10) | 4 |
| 5 | 20.51 (11.74, 28.64) | 3 | 29.56 (20.99, 37.64) | 2 | **26.98** (9.27, 44.19) | **1** | 26.86 (8.85, 42.61) | 2 |

Table S7 **Misinformation with a scientific temperament has a higher impact on drop in vaccine intent.** Weights and ranks of images in their contributions to susceptibility to misinformation about COVID-19 vaccines, where values in parentheses indicate 95% percentile intervals and values in **bold** indicate top-most/bottom-most ranked images. Top-ranked images contribute the most, while bottom-ranked the least towards drop in vaccine intent. Note that the treatment image sets differed for the UK and US, but control images are identical.

| Pre-Exposure Intent to protect self | UK | | US | |
|---|---|---|---|---|
| | **Treatment** | **Control** | **Treatment** | **Control** |
| Yes, definitely | 30.8 (28.5, 33.1) | 38.5 (34.6, 42.7) | 43.1 (40.3, 45.9) | 63.1 (58.6, 67.5) |
| Unsure, lean yes | 33.3 (30.4, 36.4) | 29.5 (24.5, 34.9) | 31.3 (28.0, 34.4) | 53.7 (47.6, 59.6) |
| Unsure, lean no | 32.1 (25.9, 38.6) | 26.4 (17.5, 36.3) | 30.8 (26.0, 35.7) | 29.7 (21.6, 38.6) |
| No, definitely not | 41.4 (34.1, 48.6) | 23.0 (11.8, 37.4) | 27.9 (23.4, 32.7) | 37.2 (28.5, 46.5) |
| **Difference to baseline: "Yes, definitely"** | | | | |
| Unsure, lean yes | 2.5 (-1.3, 6.3) | **-9.0 (-15.7, -2.5)** | **-11.8 (-16.0, -7.7)** | **-9.4 (-16.9, -2.0)** |
| Unsure, lean no | 1.3 (-5.4, 8.1) | **-12.1 (-21.8, -1.6)** | **-12.3 (-17.8, -6.5)** | **-33.4 (-42.7, -23.7)** |
| No, definitely not | **10.6 (3.0, 18.1)** | **-15.5 (-27.5, -0.5)** | **-15.1 (-20.4, -9.9)** | **-25.9 (-35.8, -15.5)** |

**Table S8 Those with lower vaccine intent had encountered more misinformation and less factual information than those with higher intent.** Values in top-half of the table indicate percentage of respondents, broken down by pre-exposure vaccine intent to protect oneself, who had recently seen images "similar" to the ones shown to them in this study—misinformation for the treatment group and factual information for the control group—for both the UK and US. Values in the bottom-half indicate probability of having seen "similar" images with respect to the baseline vaccine intent category of "yes, definitely". Values in parentheses indicate 95% percentile intervals and values in bold indicate "statistical significance" based on them.

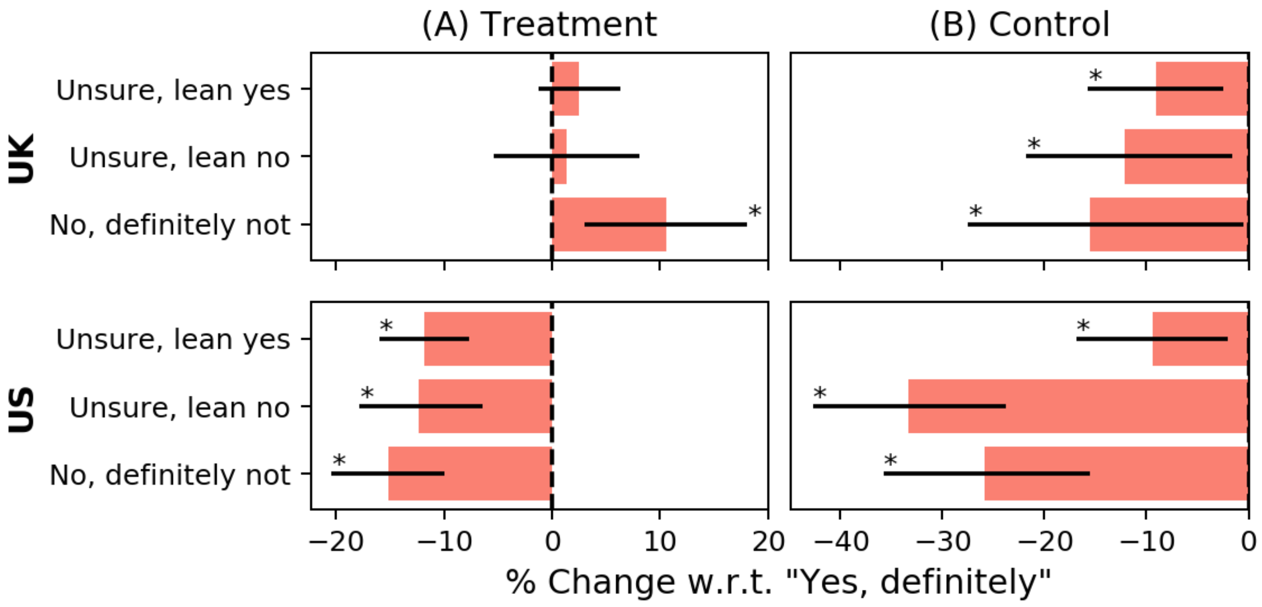


**Figure S3 Those with lower vaccine intent had encountered more misinformation and less factual information than those with higher intent.** The difference in the likelihood of respondents—broken down by pre-exposure vaccine intent to protect oneself—having seen images "similar" to the ones shown to them in this study—misinformation for the treatment group (A) and factual information for the control group (B)—for both the UK (top-row) and US (bottom-row). Black lines denote 95% percentile intervals and are starred (*) if this interval excludes zero. For example, in the UK, those who would definitely not accept the vaccine were 10.6% more likely to have encountered similar misinformation online, and 15.5% less likely to have encountered similar factual information online, when compared to those who would definitely accept the vaccine.

#### Appendix E: Questionnaire

##### Section 1: COVID-19 Knowledge Baseline

1. **[QINF]** Do you personally know anyone who has tested positive for COVID-19? If yes, was that a family member, a work colleague, a friend or someone else? (Please choose all that apply.)
   a. No
   b. Yes, myself
   c. Yes, family member in my household
   d. Yes, family member outside my household
   e. Yes, a close friend
   f. Yes, a work colleague or someone else
2. **[QSHD]** Have you been shielding because you are in a vulnerable group for coronavirus (COVID-19)?
   a. Yes
   b. No
3. **[QKNL]** How strongly do you agree or disagree with the following statements?

| Rotate Statements | Strongly agree | Tend to agree | Tend to disagree | Strongly disagree | Do not know |
|---|---|---|---|---|---|
| a) Washing hands with soap or sanitiser can help prevent the spread of coronavirus (COVID-19) | 1 | 2 | 3 | 4 | 5 |
| b) Staying indoors and reducing contact with others can help protect <u>you</u> against catching coronavirus (COVID-19) | 1 | 2 | 3 | 4 | 5 |
| c) Staying indoors and reducing contact with <u>others</u> can help protect others from catching coronavirus (COVID-19) | 1 | 2 | 3 | 4 | 5 |
| d) If you catch coronavirus (COVID-19), you can infect somebody else before you have developed symptoms | 1 | 2 | 3 | 4 | 5 |

| | | | | | |
|---|---|---|---|---|---|
| e) On average, before lockdown, someone with coronavirus (COVID-19) would have infected 2-3 other people | 1 | 2 | 3 | 4 | 5 |
| f) Treatments already exist to prevent you catching coronavirus (COVID-19) | 1 | 2 | 3 | 4 | 5 |
| g) Wearing a facemask in public can help prevent the spread of coronavirus (COVID-19) | 1 | 2 | 3 | 4 | 5 |

4. **[QCOVVCI]** How strongly do you agree or disagree with each of the following statements?

| **Rotate Statements** | **Strongly agree** | **Tend to agree** | **Tend to disagree** | **Strongly disagree** | **Do not know** |
|---|---|---|---|---|---|
| a) A coronavirus (COVID-19) vaccine would only be made available to the public if it was safe | 1 | 2 | 3 | 4 | 5 |
| b) A coronavirus (COVID-19) vaccine would only be made available to the public if it was effective | 1 | 2 | 3 | 4 | 5 |
| c) A coronavirus (COVID-19) vaccine is important | 1 | 2 | 3 | 4 | 5 |
| d) A coronavirus (COVID-19) vaccine will not be compatible with my religious or personal beliefs | 1 | 2 | 3 | 4 | 5 |
| e) I am worried that I may contract coronavirus (COVID-19) from a COVID-19 vaccine | 1 | 2 | 3 | 4 | 5 |
| f) The benefits of accepting a COVID-19 vaccine will outweigh the risks | 1 | 2 | 3 | 4 | 5 |

5. **[QVCI]** We will now ask you some questions about vaccines in general. How strongly do you agree or disagree with each of the following statements?

| Rotate Statements | Strongly agree | Tend to agree | Tend to disagree | Strongly disagree | Do not know |
|---|---|---|---|---|---|
| a) Overall, I think vaccines are important for children to have | 1 | 2 | 3 | 4 | 5 |
| b) Overall, I think vaccines are safe | 1 | 2 | 3 | 4 | 5 |
| c) Overall, I think vaccines are effective | 1 | 2 | 3 | 4 | 5 |
| d) Overall, vaccines are compatible with my religious beliefs | 1 | 2 | 3 | 4 | 5 |

##### Section 2: COVID-19 Main

6. **[QSRCUK: UK only]** What sources of information do you trust regarding COVID-19? (Please choose all that apply.)
    a. Television news
    b. Radio, podcasts and other broadcasts
    c. Newspapers and other journalism
    d. Daily government briefings
    e. National health authorities (e.g. PHE, NHS)
    f. International health authorities (e.g. WHO)
    g. Healthcare workers (e.g. doctors, nurses)
    h. Scientific experts
    i. Government websites
    j. Social media platforms (e.g. Facebook, Twitter, YouTube)
    k. Celebrities
    l. Online search engines or other websites (e.g. Google)
    m. Family and friends
    n. Work/school/college guidelines
    o. Other (specify) ________________
    p. None of the above
7. **[QSRCUS: US only]** What sources of information do you trust regarding COVID-19? (Please choose all that apply.)
    a. Television news
    b. Radio, podcasts and other broadcasts
    c. Newspapers and other journalism
    d. White House Press briefings
    e. State government briefings
    f. National health authorities (e.g. CDC)
    g. International health authorities (e.g. WHO)
    h. Healthcare workers (e.g. doctors, nurses)

i. Scientific experts
j. Government websites
k. Social media platforms (e.g. Facebook, Twitter, YouTube)
l. Celebrities
m. Online search engines or other websites (e.g. Google)
n. Family and friends
o. Work/school/college guidelines
p. Other (specify) ________________
q. None of the above

8. **[QCOVSELF]** If a new coronavirus (COVID-19) vaccine became available, would you accept the vaccine for yourself?
    a. Yes, definitely
    b. Unsure, but leaning towards yes
    c. Unsure, but leaning towards no
    d. No, definitely not
9. **[QCOVSELFWHY: if QCOVSELF!=a]** Why are you unsure about accepting a vaccine against coronavirus (COVID-19)? (Please choose all that apply.)
    a. I do not yet know enough about how safe it would be
    b. I do not yet know about how effective it would be
    c. I do not feel I am at risk of catching the virus
    d. I would want to wait until other people had been vaccinated first
    e. I do not feel I would be seriously ill if I caught the virus
    f. I am confident there will be other effective treatments soon
    g. I am confident that I have already acquired immunity (protection) through previous infection with the virus
    h. Approval/Development for the vaccine may be rushed and it may not be thoroughly tested
    i. Other, please state ________________
    j. Do not know

10. **[QCOVOTH]** If a new coronavirus (COVID-19) vaccine became available, would you accept the vaccine if it meant protecting friends, family, or at-risk groups?
    a. Yes, definitely
    b. Unsure, but leaning towards yes
    c. Unsure, but leaning towards no
    d. No, definitely not
11. **[QCOVWHEN]** When do you think the vaccine against coronavirus (COVID-19) will be publicly available for anybody to take?
    a. Less than 1 month
    b. 1 to 3 months
    c. 4 to 6 months
    d. 7 to 12 months
    e. 13 to 24 months
    f. More than 24 months
    g. I do not think a COVID-19 vaccine will ever be available
    h. Do not know

#### Section 3: Social Media

We will now ask you some questions about your use of social media

12. **[QSOCUSE]** In the past month, on average, how much time per day have you spent actively using social media?
    a. None
    b. Less than 10 minutes per day
    c. 10–30 minutes per day
    d. 31–60 minutes per day
    e. 1–2 hours per day
    f. 2–3 hours per day
    g. More than 3 hours per day
13. **[QSOCTYP]** What social media platforms do you use? (Please choose all that apply.)
    a. Facebook

   b. Twitter
   c. YouTube
   d. WhatsApp
   e. Instagram
   f. Pinterest
   g. LinkedIN
   h. Other (please state) ________________
   i. None of the above

14. **[QSOCINF]** Which of these social media platforms do you receive information regarding COVID-19 from? (Please choose all that apply.)

    *(Subset options from* **QSOCTYP***)*

15. **[QSOCSHR]** With whom do you share information regarding COVID-19? (Please choose all that apply.)
    a. Family
    b. Close friends
    c. Friends or followers on social media
    d. None of the above

16. **[QCIRSHR: if QSOCSHR!=d]** Which of these social media platforms do you share information regarding COVID-19 on?

    *(Subset options from* **QSOCTYP***)*

#### Section 4: Exposure

We will now show you 5 images, followed by a set of questions. While answering these questions, imagine these images were shared by your friends, followers or people you follow on any social media platform that you use.

*(On separate pages show images in (a) Figure 1A for UK treatments, (b) Figure 1B for US treatments, and (c) Figure 2 for controls of both countries.)*

17. **[QPOSTCOVSELF]** If a new coronavirus (COVID-19) vaccine became available, would you accept the vaccine for yourself?
    a. Yes, definitely

b. Unsure, but leaning towards yes
c. Unsure, but leaning towards no
d. No, definitely not

18. **[QPOSTCOVSELFWHY: if QPOSTCOVSELF!=a]** Why are you unsure about accepting a vaccine against Coronavirus (COVID-19)? (Please choose all that apply.)
    a. I do not yet know enough about how safe it would be
    b. I do not yet know about how effective it would be
    c. I do not feel I am at risk of catching the virus
    d. I would want to wait until other people had been vaccinated first
    e. I do not feel I would be seriously ill if I caught the virus
    f. I am confident there will be other effective treatments soon
    g. I am confident that I have already acquired immunity (protection) through previous infection with the virus
    h. Approval/Development for the vaccine may be rushed and it may not be thoroughly tested
    i. Other, please state
    j. Do not know
19. **[QPOSTCOVOTH]** If a new coronavirus (COVID-19) vaccine became available, would you accept the vaccine if it meant protecting friends, family, or at-risk groups?
    a. Yes, definitely
    b. Unsure, but leaning towards yes
    c. Unsure, but leaning towards no
    d. No, definitely not

We will now ask you questions about each image that you were shown.

*(Loop over every image X = {1, 2, 3, 4, 5} re-shown followed by this set of questions.)*

20. **[QPOSTVACX]** Overall, the information provided in this image makes me
    a. Much less inclined to be vaccinated
    b. A little less inclined to be vaccinated
    c. No less or more inclined to be vaccinated
    d. A little more inclined to be vaccinated

e. Much more inclined to be vaccinated

f. Do not know

21. **[QPOSTBELIEFX]** Overall, how much do you agree with the information in this image?

a. Strongly agree

b. Somewhat agree

c. Neither agree nor disagree

d. Somewhat disagree

e. Strongly disagree

f. Do not know

22. **[QPOSTTRUSTX]** Overall, how much do you think this information is trustworthy?

a. Very trustworthy

b. Somewhat trustworthy

c. Neither trustworthy nor untrustworthy

d. Somewhat untrustworthy

e. Very untrustworthy

f. Do not know

23. **[QPOSTCHECKX]** Overall, how likely are you to fact-check the information in this image via other sources?

a. Very likely

b. Somewhat likely

c. Neither likely nor unlikely

d. Somewhat unlikely

e. Very unlikely

f. Do not know

24. **[QPOSTSHARE]** Overall, how likely are you to share this image with your friends or followers?

a. Very likely

b. Somewhat likely

c. Neither likely nor unlikely

d. Somewhat unlikely

e. Very unlikely

f. Do not know

*(End loop.)*

#### Section 5: Information sharing

Taking into account the five images we showed you, please answer the following questions

25. **[QPOSTSIM]** Have you seen similar content online in the last month on social media?

    a. Yes

    b. No

    c. Do not know

26. **[QPOSTFRQ: if QPOSTSIM=a]** How often have you seen similar content being shared on social media in the last one month?

    a. Multiple times a day

    b. Once or twice a day

    c. A few times a week

    d. A few times a month

    e. Never

    f. Do not know

27. **[Q31b: if QPOSTSIM=a]** Have you shared, liked, or commented on similar content in the last month?

    a. Yes

    b. No

    c. Do not know

#### Section 6: Impact of COVID-19

28. **[QCOVAFF]** How strongly do you agree or disagree with each of the following statements?

| Rotate Statements | Strongly agree | Tend to agree | Tend to disagree | Strongly disagree | Do not know |
|---|---|---|---|---|---|
| a) COVID-19 has negatively impacted my mental health and wellbeing in the last 6 months | 1 | 2 | 3 | 4 | 5 |

| | | | | | |
|---|---|---|---|---|---|
| b) COVID-19 has negatively impacted my financial stability over the last 6 months | 1 | 2 | 3 | 4 | 5 |
| c) COVID-19 has severely disturbed my daily life over the last 6 months | 1 | 2 | 3 | 4 | 5 |
| d) COVID-19 has hampered my ability to socialize with family and close friends over the last 6 months | 1 | 2 | 3 | 4 | 5 |

**Section 7: Demographics**

We will now ask you some questions about yourself

29. **[DGEOUK: UK only]** Please select your region and local authority

    *(Display drill-down menu for UK regions and local authorities.)*

30. **[DGEOUS: US only]** Please select your state and county

    *(Display drill-down menu for US states and counties.)*

31. **[DAGE]** What is your age?

    *(Numeric values from 18 to 120.)*

32. **[DGEN]** What is your gender?
    a. Male
    b. Female
    c. Other
    d. Prefer not to answer

33. **[DEDUUK: UK only]** What is the highest level of education you have completed? If currently enrolled, mark the highest qualification received.
    a. No academic or professional qualifications
    b. 0-4 GCSE, O-level or equivalents
    c. 5+ GCSE, O-level, 1 A level, or equivalents
    d. Apprenticeship
    e. 2+ A levels, or equivalents
    f. Undergraduate degree

g. Postgraduate degree or other professional degrees
h. Other (e.g. vocational, foreign qualifications):________________
i. Prefer not to answer

34. **[DEDUUS: US only]** What is the highest level of education you have completed? If currently enrolled, mark the previous grade or highest qualification received.
    a. No academic or professional qualifications
    b. Nursery or preschool through grade 12
    c. High school diploma or GED
    d. 2-year college degree
    e. 4-year college degree
    f. Postgraduate degree or other professional degrees
    g. Other: ________________
    h. Prefer not to answer
35. **[DEMP]** Which of the following best describes your working status <u>6 months ago</u>?
    a. Working full-time (include self-employed)
    b. Working part-time (include self-employed)
    c. Unemployed
    d. Student
    e. Looking after family or home
    f. Retired
    g. Long-term sick or disabled
    h. Prefer not to answer
36. **[DREL]** How would you describe your religious affiliation?
    a. Roman Catholic
    b. Protestant
    c. Other Christian: ________________
    d. Jewish
    e. Hindu
    f. Muslim

g. Buddhist

h. Other: ________________

i. Atheist or agnostic

j. Prefer not to answer

37. **[DPOLUK: UK only]** Generally speaking, do you consider yourself as

    a. Conservative

    b. Labour

    c. Liberal Democrat

    d. SNP

    e. Other: ________________

    f. Don't know

    g. Prefer not to answer

38. **[DPOLUS: US only]** Generally speaking, how would you describe your political affiliation?

    a. Republican

    b. Democrat

    c. Independent

    d. Don't know

    e. Prefer not to answer

39. **[DPOLUSIND: US only, if DPOLUS=c or DPOLUS=d]** As of today, do you politically lean more towards

    a. Republican Party

    b. Democratic Party

    c. Don't know

    d. Prefer not to answer

40. **[DETHUK: UK only]** Which best describes your ethnicity? (Please choose one response that best applies.)

    a. White: English/Welsh/Scottish/Northern Irish/British

    b. White: Irish

    c. White: Other white background

d. White and Black Caribbean, or White and Black African
e. White and Asian
f. Asian or Asian British: Indian
g. Asian or Asian British: Pakistani
h. Asian or Asian British: Bangladeshi
i. Asian or Asian British: Chinese
j. Asian or Asian British: Other
k. Black, African, Caribbean, or Black British
l. Other: ________________
m. Prefer not to answer

41. **[DETHUS: US only]** Which best describes your ethnicity? (Please choose one response that best applies.)
    a. Non-Hispanic White
    b. Hispanic
    c. Black or African American
    d. American Indian or Alaska Native
    e. Asian
    f. Native Hawaiian or Pacific Islander
    g. Other: ________________
    h. Prefer not to answer
42. **[DLANUK: UK only]** What is your main language?
    a. English
    b. Polish
    c. Punjabi
    d. Urdu
    e. Bengali
    f. Other: ________________
    g. Prefer not to answer
43. **[DLANUS: US only]** What is your main language?

a. English

b. Spanish

c. Chinese

d. French

e. Other: ________________

f. Prefer not to answer

44. **[DINCUK: UK only]** What is your total annual household income in GBP (£) from all sources before tax?

a. Under £15,000

b. £15,000 - £24,999

c. £25,000 - £34,999

d. £35,000 - £44,999

e. £45,000 - £54,999

f. £55,000 - £64,999

g. £65,000 - £74,999

h. £75,000 - £84,999

i. £85,000 - £94,999

j. £95,000 or over

k. Prefer not to answer

45. **[DINCUS: US only]** What is your total annual household income in USD ($) from all sources before tax?

a. Under $15,000

b. $15,000 - $24,999

c. $25,000 - $34,999

d. $35,000 - $44,999

e. $45,000 - $54,999

f. $55,000 - $64,999

g. $65,000 - $74,999

h. $75,000 - $84,999

i. $85,000 - $94,999

j. $95,000 or over

k. Prefer not to answer

**Section 8: Debrief**

The aim of this study was to monitor your perceptions towards a COVID-19 vaccine and to assess whether the images we showed you changed your perceptions towards vaccinating. The images we showed you are all examples of online information that contains either misleading or incorrect information about a COVID-19 vaccine.

**[UK only]**

For up-to-date information surrounding the COVID-19 pandemic, please consult the NHS's coronavirus webpage https://www.nhs.uk/conditions/coronavirus-covid-19

**[US only]**

For up-to-date information surrounding the COVID-19 pandemic, please consult the coronavirus webpage at the US Centers for Disease Control https://www.cdc.gov/coronavirus/2019-ncov/index.html
